# SARS-CoV-2 viral replication persists in the human lung for several weeks after symptom onset

**DOI:** 10.1101/2023.03.06.23286834

**Authors:** M Tomasicchio, S Jaumdally, L Wilson, A Kotze, L Semple, S Meier, A Pooran, A Esmail, K Pillay, R Roberts, R Kriel, R Meldau, S Oelofse, C Mandviwala, J Burns, R Londt, M Davids, C van der Merwe, Roomaney A, L Kühn, T Perumal, A.J Scott, M.J Hale, V Baillie, S Mahtab, C Williamson, R Joseph, A Sigal, I Joubert, J Piercy, D Thomson, DL Fredericks, MGA Miller, M.C Nunes, S.A Madhi, K Dheda

## Abstract

**Rationale:** In the upper respiratory tract replicating (culturable) SARS-CoV-2 is recoverable for ∼ 4 to 8 days after symptom onset, however, there is paucity of data about the frequency or duration of replicating virus in the lower respiratory tract (i.e. the human lung).

**Objectives:** We undertook lung tissue sampling (needle biopsy), shortly after death, in 42 mechanically ventilated decedents during the Beta and Delta waves. An independent group of 18 ambulatory patents served as a control group.

**Methods:** Lung biopsy cores from decedents underwent viral culture, histopathological analysis, electron microscopy, transcriptomic profiling and immunohistochemistry.

**Results:** 38% (16/42) of mechanically ventilated decedents had culturable virus in the lung for a median of 15 days (persisting for up to 4 weeks) after symptom onset. Lung viral culture positivity was not associated with comorbidities or steroid use. Delta but not Beta variant lung culture positivity was associated with accelerated death and secondary bacterial infection (p<0.05). Nasopharyngeal culture was negative in 23.1% (6/26) of decedents despite lung culture positivity. This, hitherto, undescribed bio-phenotype of lung-specific persisting viral replication was associated with an enhanced transcriptomic pulmonary pro-inflammatory response but with concurrent viral culture positivity.

**Conclusions:** Concurrent, rather than sequential active viral replication continues to drive a heightened pro-inflammatory response in the human lung beyond the second week of illness and was associated with variant-specific increased mortality and morbidity. These findings have potential implications for the design of interventional strategies and clinical management of patients with severe COVID-19 disease.

**At a Glance Commentary:** *Scientific Knowledge on the Subject:* Investigations to understand SARS-CoV-2 viral shedding (determined by PCR or antigen testing) have extensively focused on samples from the upper respiratory tract. The widely accepted view is that acute severe SARS-CoV-2 infection is characterised by a viral replicative phase in the first week of symptomatic illness followed by a pro-inflammatory immunopathologic phase peaking in the second and third weeks of illness. However, it remains unclear whether detection of SARS-CoV-2 beyond 2 weeks after symptom onset in published studies represent active replication competent virus because it may represent residual genomic or antigenic material in the tissue.

*What This Study Adds to the Field:* We have identified a, hitherto, undescribed bio-phenotype of acute severe COVID-19 characterised by persisting viral replication in the lung for up to 4 weeks after symptom onset. ∼40% of acute severe COVID-19 intensive care unit (ICU) decedents (n=42) had nasopharyngeal swab culture positivity at ∼2 weeks post-symptom onset versus only ∼5% in a group of ambulatory control patients (n=18). There was compartment-specific (nasopharynx versus lung) discordance. The phenotype of lung-specific persisting viral replication was associated with variant-specific accelerated death, an exaggerated inflammatory response, and attenuated T-cell immunity in the lung (based on histopathological and transcriptomic studies). This challenges the traditional view that viral replication occurs during the first 5 to 10 days of illness, which is followed by an effector or hyperinflammatory phase. This is the first study, to our knowledge, to systematically culture virus from the human lung and map out its related clinical determinants, and which describes the human lung transcriptomic profile of culture-positive versus culture-negative patients with severe COVID-19 disease.

## Introduction

Coronavirus disease-19 (COVID-19) caused by the Severe Acute Respiratory Syndrome Coronavirus-2 (SARS-CoV-2) has been the foremost killer globally over the last 3 years. Case fatality risk in hospitalised patients, and particularly in mechanically ventilated patients, during the Beta and Delta waves was particularly high [∼50%-70%; (1)]. Even with the Omicron-related variants, case fatality risk remains significant in the elderly and immunocompromised persons, and in several countries including the UK, Italy, France, Brazil, and prominently in China where there is now an ongoing epidemic of severe COVID-19 disease (2–10). Better therapeutic interventions are needed. However, despite considerable research, the pathogenesis of severe COVID-19, relative to viral kinetics, remains incompletely understood.

SARS-CoV-2 detection (ascertained through PCR positivity or antigen detection) can persist for several weeks from symptom onset (11). Post-mortem studies have shown persistence of SARS-CoV-2 in tissues detected by PCR and immunohistochemistry for up to several weeks after symptom onset (12, 13). However, detection of SARS-CoV-2 in these studies may not represent replication competent virus (detectable only by viral culture) but residual genomic or antigenic material in the tissues. Shedding of replicating virus confirmed through serial viral culture (i.e. *in vitro* replication in human cell lines) from the upper respiratory tract (URT) has been shown to persist for only ∼2 to 8 days after symptom onset (11, 14–23). These findings have been confirmed in human lung challenge studies with viable pathogen where virus was cultured from the URT until a median of 4 days (and a maximum of 10 days) from symptom onset (24). However, hardly anything is known about the compartment-specific duration of actively replicating virus in the lower respiratory tract (LRT), particularly in acute severely ill hospitalised patients undergoing mechanical ventilation. We hypothesised that there is compartment-specific uncoupling of viral replication in severe COVID-19 i.e. replicating virus can persist in the LRT beyond 10 days from symptoms onset, independent of its persistence in the URT, and this persistence may be associated with an altered pulmonary immunity.

## Methods

### Patients

The decedents (n=42) were recruited from Chris Hani Baragwanath Academic Hospital, Johannesburg, South Africa (n=18; Beta group) and Groote Schuur Hospital, Cape Town, South Africa (n=24; Delta group). Figure 1A outlines an overview of the study plan. Ambulatory controls (n=18) were recruited at diagnosis (baseline; ∼5 days from symptom onset), 7 days and 14 days post diagnosis. Minimally invasive tissue samples (MITS) and nasopharyngeal swabs from decedents (n=42) in the Beta and Delta waves (Figure 1B) were taken immediately after death. In addition, heart, liver, kidney, and adipose tissue samples were also taken from the Delta variant decedent cohort only. Ethical approval was obtained from the Human Research Ethics Committee (HREC) of the University of Cape Town (HREC approval number 866/2020) and University of Witwatersrand (HREC approval number M200313). Biosafety approvals were obtained from the Faculty Biosafety Committee of the University of Cape Town (IBC008-2021).

**Figure 1.**
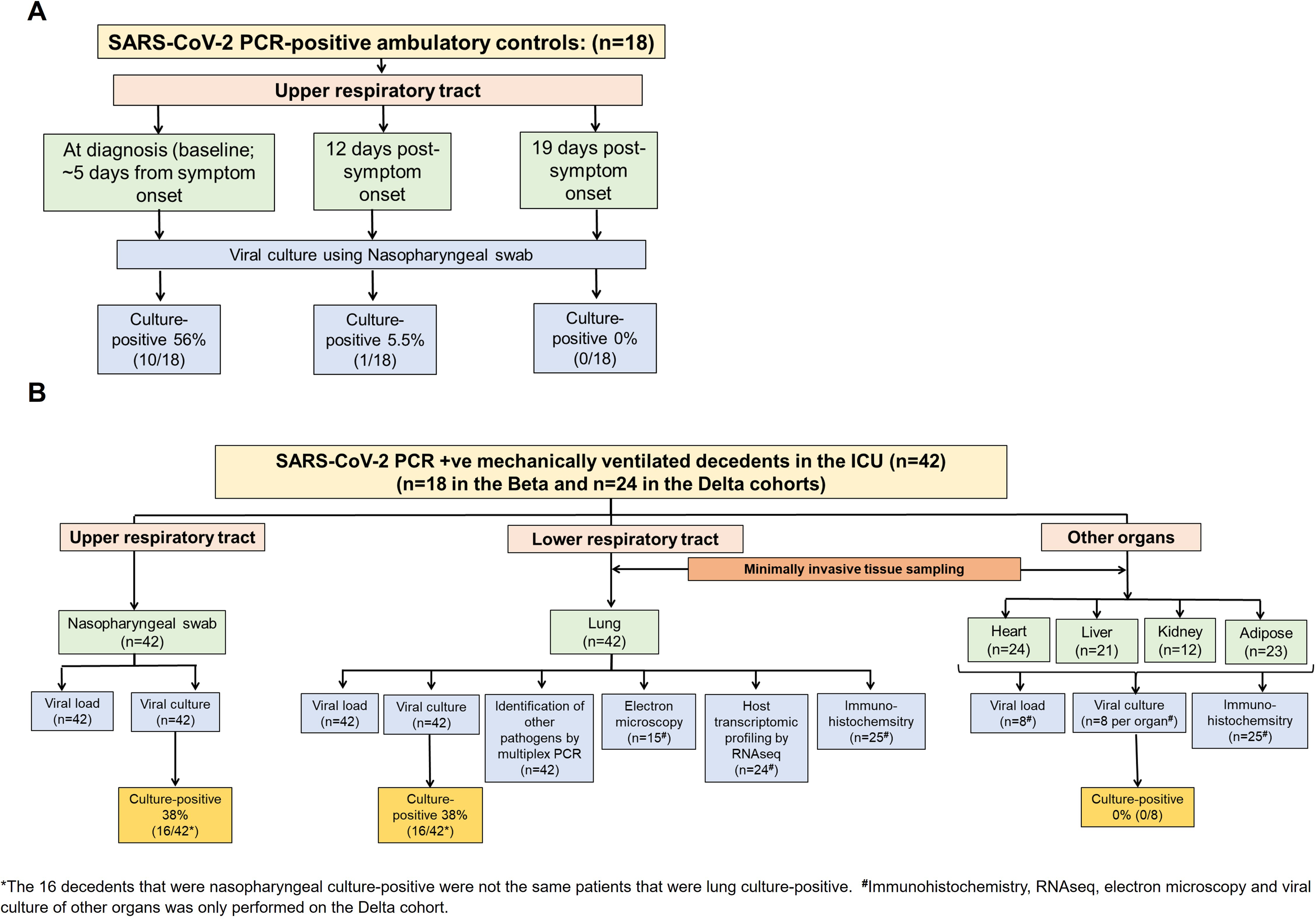
Study overview including SARS-CoV-2 PCR-positive ambulatory controls (A) and mechanically ventilated decedents (B) recruited during the Beta and Delta waves. Nasopharyngeal (NP) swabs from ambulatory COVID-19 controls were obtained approximately 5 days after symptom onset (diagnosis), and then at 12- and 19-days post symptom onset. Minimally invasive tissue samples (MITS) and NP swabs were retrieved from decedents shortly after death.

### Viral culture

To establish the *in vitro* viral culture model, a SARS-CoV-2 viral stock was used to infect the human lung carcinoma cell line, H1299 ACE2, in a BSL3 laboratory and infection was confirmed by light microscopy (as assessed by cytopathic effects of the virus on the cell line) and confocal microscopy (Figure S1A and B). Serial dilutions of the viral stock were used to establish the limit of detection of the PCR assay at 1×10^1^ copies/ml (Figure S1C). Viral culture was performed on the nasopharyngeal swab and lung biopsy samples as indicated in the study overview (Figure 1) and detailed in the online supplement. Viral culture result reproducibility was good (see online supplement).

### Multiplex PCR to detect secondary bacterial infections

The lung biopsy cores, stored in universal transport medium, were briefly homogenised and 200µl of the supernatant was applied to the BioFire FilmArray Pneumonia panel (Bioméieux, South Africa). The panel was run using protocol BAL v3.3 according to the manufacturer’s instructions, thus generating RT-PCR readouts for 33 bacterial and viral pathogens. Bronchopneumonia was defined as histological evidence of a neutrophilic alveolar infiltration together with the detection of bacterial genomic material in the biopsy cores.

### Immunohistochemistry

Immunohistochemical staining was performed using the Roche Ventana Automated platform (Ventana XT autostainer) as indicated by the manufacturer. Tissue sections were prepared, stained, and viewed using standard techniques (25). Antibodies included anti-CD3 (2GV6), and anti-CD8 (SP57) (Roche USA).

### Haematoxylin & eosin (H&E) staining and transmission electron microscopy (TEM)

H&E staining and TEM were performed according to standard procedures (25). H&E-stained slides were viewed using an Olympus BX43 microscope. TEM tissue sections were viewed using a Carl Zeiss EM109 microscope.

### SARS-CoV-2 whole genome sequencing

Total SARS-CoV-2 RNA was extracted from lung biopsy samples and whole genome sequencing was performed. The generated reads were analysed with the Exatype (https://exatype.com) software to identify minor and major variants. The assembled consensus sequences were analysed using Nextclade Web (https://clades.nextstrain.org) for further quality control and clade assignment.

### RNAseq

Total RNA was extracted from lung biopsy samples from the Delta group, sequenced and mapped consecutively to the human and COVID reference genomes using the Spliced Transcripts Alignment to a Reference (STAR) software [version 2.7.7a, (26)]. A differential expression (DE) analysis was performed on the generated raw read count file with the edgeR (Version 3.38.4) R package (27). The DE results were ranked by fold change and the gseGO function, from the clusterProfiler R clusterProfiler [Version 4.0, (28)] R package was used to perform a gene set enrichment analysis (GSEA) for the Gene Ontology Biological Process pathways. Pathways with an FDR <0.05 were considered significant.

### Confocal microscopy

The H1299 ACE2 cells were plated, infected with SARS-CoV-2 and allowed to adhere to coverslips slides overnight at 37°C. The next day the cells were stained with or without anti-SARS-CoV-2 S1 spike protein (ThermoFisher, USA) and the slides were mounted in Mowiol (Calbiochem, USA) containing n-propyl gallate (Sigma-Aldrich, Germany) as an anti-fading agent. Confocal microscopy was performed with a Zeiss Axiovert 200M LSM510 Meta NLO Confocal Microscope.

### Sample size calculation and statistical analysis

We hypothesised that we would detect lung culture positivity at 14 days post-symptom onset in ∼33% of decedents. A sample size of ∼40 participants would allow us to ascertain that level of positivity with a 15% margin of error using 95% confidence and 80% power (OpenEpi, Version 3, opensource calculator).

The Fisher Exact test was employed for categorical variables and for continuous variables, Mann-Whitney test was used for non-parametrically distributed data between the culture-negative and culture-positive groups (Stata version 17 or GraphPad, Version 9.4.1). A p-value of < 0.05 was considered significant for all statistical analyses.

The multivariable analysis was performed in R by fitting a binomial Generalized Linear Model (GLM) to assess the association between steroid use and the presence of secondary bacterial infection on culture status. The tidymodels (version, 1.0.0) R package was used to perform predictive modelling using the glm binomial classification algorithm. To account for the small sample size, 1000 bootstraps were performed for each analysis using the “bootstraps” function (non-parametric) from the rsample package (version1.2.0).

## Results

### Demographics and clinical characteristics of the decedents

The demographics of patients enrolled in the study are shown in Table S1. The median age of the patients was 53 years with 48% being males (20/42). 40.5% (17/42) had a secondary bacterial infection and 11% (4/38) had bacterial bronchopneumonia (microbiologically and histopathological confirmed). The median time from onset of symptoms to death, ICU admission to death and high flow oxygen admission to death was 17 (IQR; 9-22), 5 (2–12) and 11 (6–15) days, respectively.

### SARS-CoV-2 replicating persistence in the human lung of mechanically ventilated decedents

We first ascertained the frequency and duration of replicating virus in lung tissue (which to our knowledge has not been previously undertaken). Culturable virus in the lung was present in 38.1% (16/42; Figure 2A) of mechanically ventilated ICU decedents, at a median of 15 days (and up ∼4 weeks; Figure S2) from symptom onset to sampling/death (Figure 3A). As expected, 56% (10/18) of a prospectively recruited control group of ambulatory patients had culturable virus, using nasopharyngeal swab samples, at day 5 from symptom onset (Figure 2B). In the same group of patients after 12- and 19- days after symptom onset, only 5.5% (1/18) and 0% (0/18), respectively, had culturable virus from their nasopharyngeal swab (Figure 2B). By contrast, 38% of nasopharyngeal swabs from the mechanically ventilated ICU descendants had culturable virus (Figure 2B), at a median of 13 days from symptom onset to sampling/death (Figure S3B). Additionally, SARS-CoV-2 could be detected by PCR in multiple organs in lung culture-positive decedents in the Delta cohort (biopsies other than the lung was not performed in the Beta cohort) suggesting widespread multi-organ viral dissemination (Figure 2C). SARS-CoV-2 was also detected in adipose tissue of culture-positive decedents (hitherto undescribed). We did not culture virus from the organs of the decedents other than the lung. Thus, viral genetic material was only detectable by PCR in the lung culture-positive patients in the other organs. This probably indicates that the virus disseminated systemically in these patients, who had chronic replicative disease in the lung and not in the other organs. We therefore only presented the PCR results from the other organs for the lung-culture positive patient samples in Figure 2C. Clinical characteristics, such as, age and comorbidities were similar in the lung culture-positive versus culture-negative groups (Table S1). We found no association between viral genetic variant and the phenotype of replicating viral persistence (although this might have been a factor of the limited sample size; Table S5).

**Figure 2.**
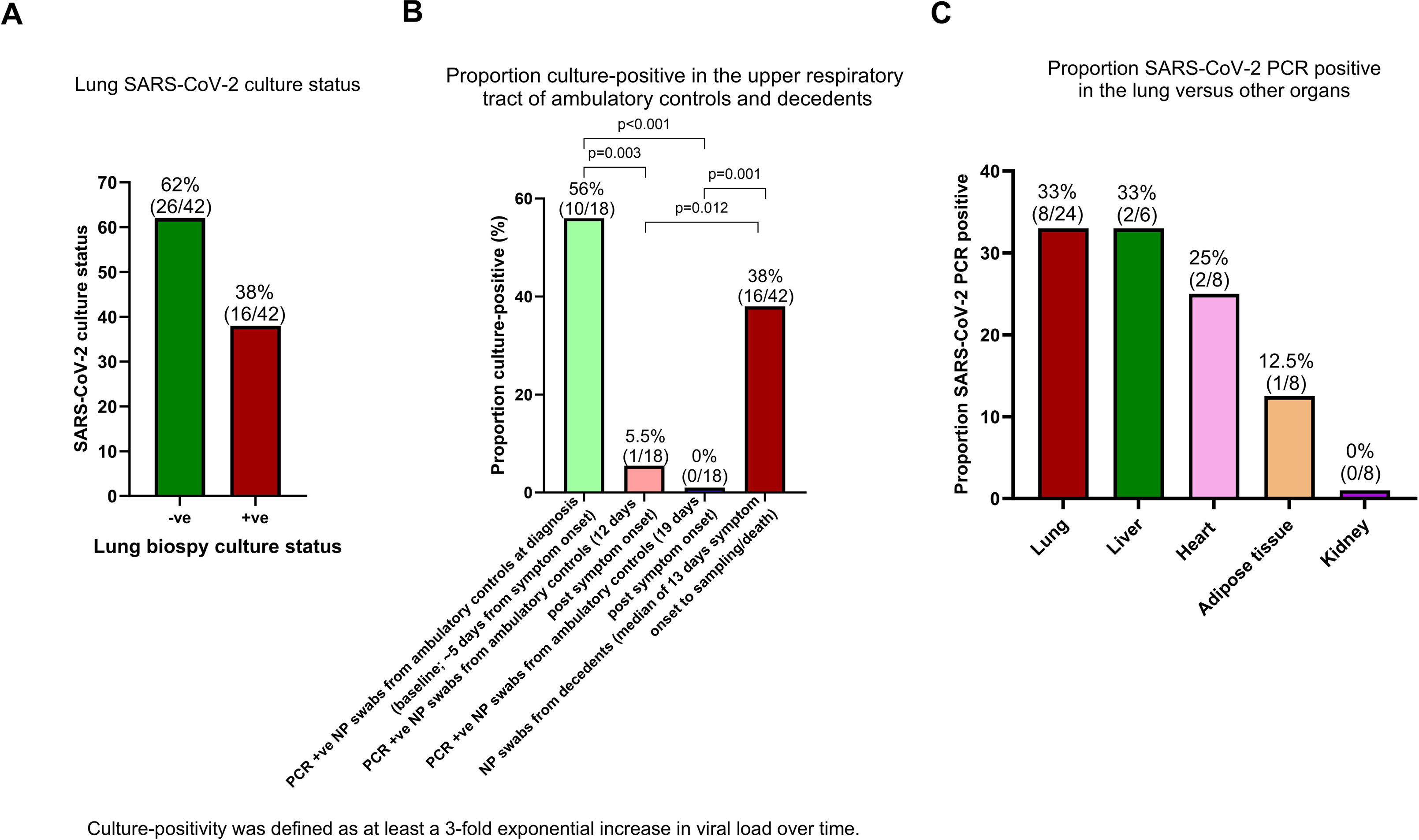
Active replicating virus was recovered from the lungs of over one third of decedents (16/42). **(A)** Proportion of lung biopsy samples that were culture-positive from the decedents. **(B)** Proportion of ambulatory patient and decedent NP swab samples that were culture-positive. **(C)** PCR positivity of organs of lung culture-positive decedents from the Delta cohort (organs other than the lung were not culture positive). NP= nasopharyngeal.

**Figure 3.**
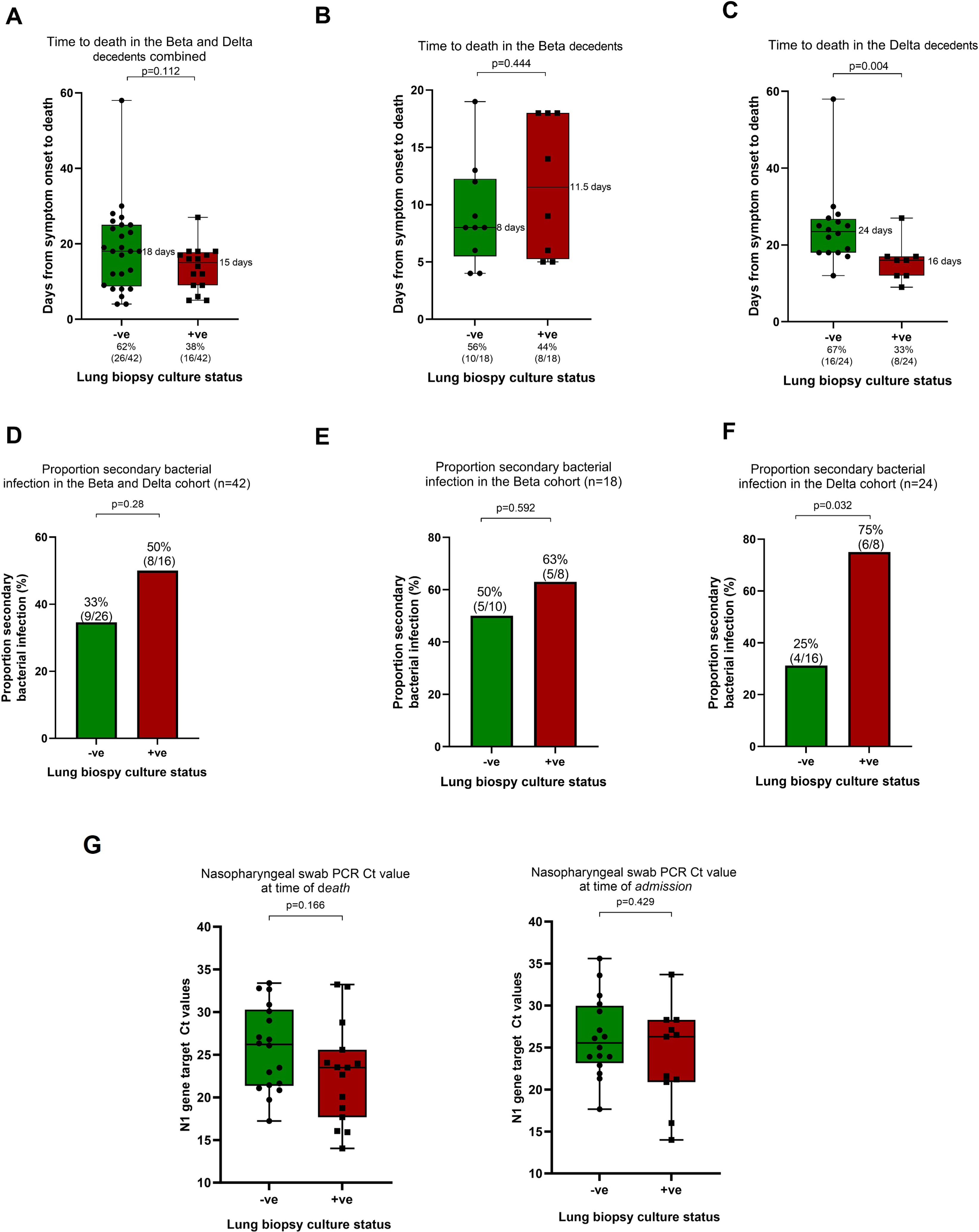
The phenotype of replicating viral persistence, compared to the culture-negative participants, was associated with accelerated death and a higher frequency of bacterial bronchopneumonia in the Delta but not the Beta group. (**A**) The days from symptom onset to death for the culture-negative (-ve; green) and culture-positive (+ve; red) groups for both groups combined, and for the Beta (**B**) and Delta (**C**) groups alone. Proportion of samples/participants with a secondary bacterial infection in culture-negative and culture-positive decedents overall i.e. the combined groups (Beta and Delta) (**D**), Beta group only (**E**), and Delta group only (**F**). **(G)** PCR cycle threshold (Ct) value at the time of death or at admission could not discriminate or predict lung culture status. The nasopharyngeal swab PCR Ct values at admission or death were missing for some participants because they were either diagnostically confirmed by antigen testing or the Ct value was not recorded. Due to the nature of the pandemic and the burden of the disease on the healthcare infrastructure at the time, Ct values at peak periods were not recorded. We have conducted sensitivity and imputation analyses indicating that these missing data points are redundant.

### Time to death in the Delta and Beta groups and predictors of lung culture **positivity.**

Next, we evaluated variant-specific relationships to clinical outcomes. Mechanically ventilated patients who were SARS-CoV-2 lung culture-positive in the Delta, but not the Beta group, had a higher proportion of accelerated death (i.e. shorter duration from symptom onset to death; Figure 3C versus 3B; p=0.004), and a higher proportion of lung-specific secondary bacterial infection (Figure 3F versus 3E; p=0.032) compared to culture-negative decedents. Similarly, to the lung culture data, the nasopharyngeal swab culture-positive Delta, but not the Beta group, had a higher proportion of accelerated death (Figure S3D versus S3C; p=0.026).

The bacterial species identified from the lung biopsies of both the Beta and Delta groups included *Streptococcus, Staphylococcus, Haemophilus, Acinetobacter*, Proteus spp, *Escherichia, Klebsiella, Enterobacter* and *Serratia* (Table S4). Overall, both groups were infected with one or more bacteria that were sensitive or resistant to β-lactams and/or carbapenems (Table S4). Key clinical and demographic characteristics such as differences in co-morbidities (age, obesity, diabetes, HIV positivity etc; Table S1) associated as drivers of severe COVID-19 disease and poor prognosis, could not explain these observations, despite the lower population-level vaccination and pre-existing COVID-19 exposure rates in the Beta cohort. Steroid usage (proportion) was similar in the culture-positive and culture-negative groups (Table S1; though the duration of steroid usage was significantly higher in the culture negative group), and there was no significant (p>0.05) association between steroid use and lung culture positivity or the presence of secondary bacterial infection in a multivariable analysis. If anything, there was a trend (p=0.06) to greater steroid exposure in the lung culture negative group (in the multivariable analysis) arguing against its role in driving viral replication.

Next, we interrogated whether nasopharyngeal PCR characteristics (Ct value), either at admission or close to death, could identify the phenotype of lung replicating viral persistence. However, nasopharyngeal Ct neither at admission, nor at the time of death was associated with lung culture positivity (Figure 3G). This suggests that the kinetics of viral replication was different in the upper and the lower respiratory tract.

### Lung immunity and histology of the culture-negative versus culture-positive groups

We then ascertained whether the phenotype of replicating viral persistence was associated with attenuated or modulated lung immunity in the Delta decedents (transcriptomic and flow cytometric studies were only carried out in Cape Town, i.e the Delta decedents, due to location-specific availability of assays and limited Beta group biopsy cores that had been used for unrelated studies). Immunohistochemical staining indicated that there was significantly less infiltration of CD3+ T-cells, specifically CD8+ T-cells in the alveoli and interstitium of the SARS-CoV-2 culture-positive compared with the culture-negative individuals in the Delta decedents (Figure 4A and B).

**Figure 4.**
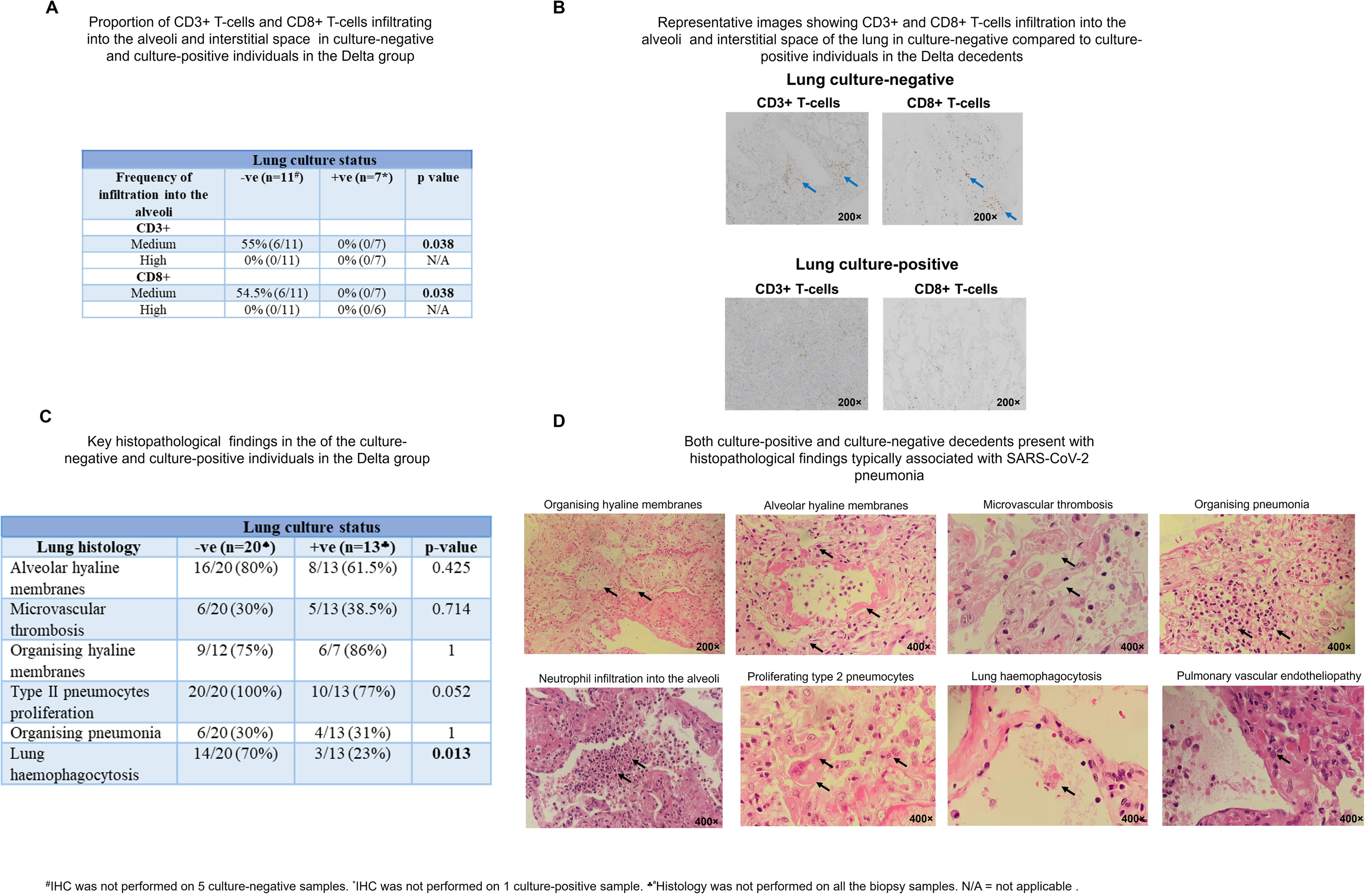
A higher proportion of T-cells, macrophages and pneumonocytes infiltrate into the lung of the culture-negative versus culture-positive decedents in the Delta group. **(A)** More CD3+ and CD8+ T-cells infiltrate into the alveoli and interstitial space of the lung culture-negative versus culture-positive group in the Delta decedents as assessed by immunohistochemistry. **(B)** Representative images (immunohistochemistry) at 200× magnification showing increased T-cell infiltration into the interstitial space (blue arrow) in the lung culture-negatives versus the culture-positives in the Delta cohort. The density of CD3+ or CD8+ T-cells in the alveoli or interstitial tissue were assessed and scored as medium or high. A magnitude of 10%-50% or >50% was defined as medium or high infiltration, respectively. Histopathology findings **(C)** and representative images **(D)** associated with diffuse alveolar damage and microvascular thrombosis in the Delta decedents. The black arrows indicate key histopathological features. The magnification settings were either set at 200× or 400×. The relative magnification of each light microscopy image is shown.

The typical histological features of severe COVID-19 (e.g. diffuse alveolar damage and microvascular thrombosis) were similar in the SARS-CoV-2 culture-positive and the culture-negative phenotype suggesting that these events occurred in the early rather than the persistent viral replication phase (Figure 4C, D and S4; Table S2, S3). Interestingly, we observed that some features of leukocyte hyperactivation (i.e., hemophagocytic syndrome) were more common in the SARS-CoV-2 culture-negative versus the culture-positive group, potentially in keeping with an aberrant immune response characterised by a lack of immune regulation, as outlined above (Figure 4C and 4D; p=0.013).

The transcriptional analysis of post-mortem lung tissue after adjustment for multiple testing, identified a total of 11 up- and 4 down-regulated genes in the culture-positive versus culture-negative groups (FDR<0.05; specific genes discussed further in the online supplement; Figure S6). To ensure that the transcriptional signal was uniform, lung biopsy cores from each decedent were placed in 1 tube containing RNAlater to ensure that enough genetic material was obtained. The lung-culture-positive group expressed higher levels of carbonic anhydrase 12 (CA12) than the lung culture-negative group (Figure S6 and Table S6). This protein induces a phenotype similar to high-altitude pulmonary oedema with a decreased ratio of arterial oxygen, partial pressure to fractional inspired oxygen, and a reduction of the carbon dioxide levels (29). This was associated with increased tachypnoea and fibrinogen levels/fibrin formation and the presence of hypoxia leading to acute respiratory distress syndrome [ARDS; (29)]. Another gene that was highly overexpressed in the culture-positive cohort was CD177, a glycosylphosphatidylinositol (GPI)-anchored protein expressed by neutrophils. CD177 plays a key role in neutrophil activation, transmigration and adhesion to the endothelium and is associated with the severity of COVID-19 disease (Figure 6, S6 and Table S6) (30). Fu et al (31) reported a high neutrophil to lymphocyte ratio in the alveolar spaces of the lung from deceased patients with COVID-19. Elevated levels of CD177 were recently identified by transcriptomics in the peripheral blood (32) and by proteomics in bronchoalveolar lavage cells (33) of COVID-19 patients with mild and severe disease, which supports our data of an upregulation of CD177 in the lung culture-positive decedents.

Syndecan binding protein 2 was significantly upregulated in the culture-positive versus the culture-negative group (Figure S6 and Table S6). The protein is a family member of the syndecans (SDC) which are transmembrane proteoglycans that facilitate the cellular entry of SARS-CoV-2 (34). Endothelial cells express SDC2 and during virus internalization and syndecans colocalize with ACE2, suggesting a jointly shared internalization pathway. Hudak et al (34) reported that entry via SDCs enabled efficient gene transduction with SARS-CoV-2 pseudovirus which implied that SDC-mediated internalisation pathway maintained the viral particles biological activity. Viruses that target SDCs in the lung may therefore interfere with SDC-dependent signalling as inhibitors to both ACE2 and syndecan reduced the cellular entry of SARS-CoV-2, thus supporting the complex nature of internalization.

The GSEA performed using the full list of differentially expressed genes ranked by fold-change, identified activated pathways that were associated with a proinflammatory response related to cytokine signalling, neutrophil and monocyte chemotaxis/recruitment, and viral entry/defence, all of which are implicated in COVID-19-related hypercytokinaemia (35) (Figure 5, 6, S5, S6, Table S6 and S7A). Significantly repressed pathways were generally associated with body homeostasis (Figure 5A and B). There was also in tandem upregulation of Th1 and Th17 signalling pathways (Table S7B) but to a substantially lesser extent than that of innate cellular and signalling pathways (IL-1, IL-6 and neutrophil-related; Table S7A). These features may be consistent with an aberrant immune response including a lack of activation of regulatory and immune-suppressive pathways. T-cell exhaustion consistent with upregulation of PD-1, CTLA-4 and LAG (Table S7C) known to be associated with severe COVID-19, was not observed.

**Figure 5.**
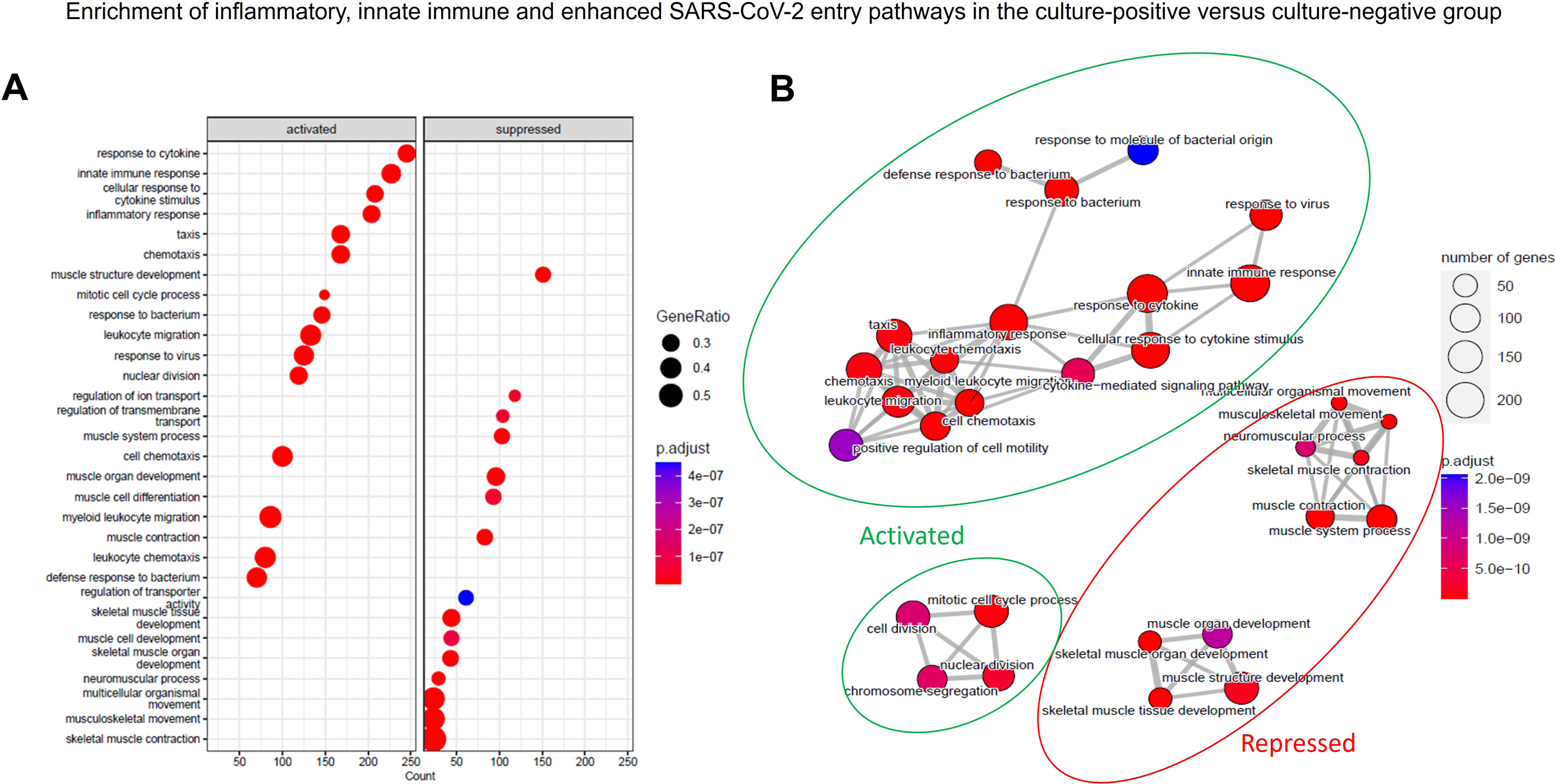
The transcriptomic analysis revealed that the culture-positive group, in comparison to the culture-negative group, had enrichment of activated pathways associated with inflammation, innate immunity, responses to cytokines, and responses to virus/ bacterial stimuli in the Delta descendants. Dot plot illustrating the significantly activated and suppressed pathways along with the gene count and ratio for each pathway (**A**), enrichment map illustrating the significantly activated and suppressed pathways along with the gene count and ratio for each pathway (**B**).

**Figure 6.**
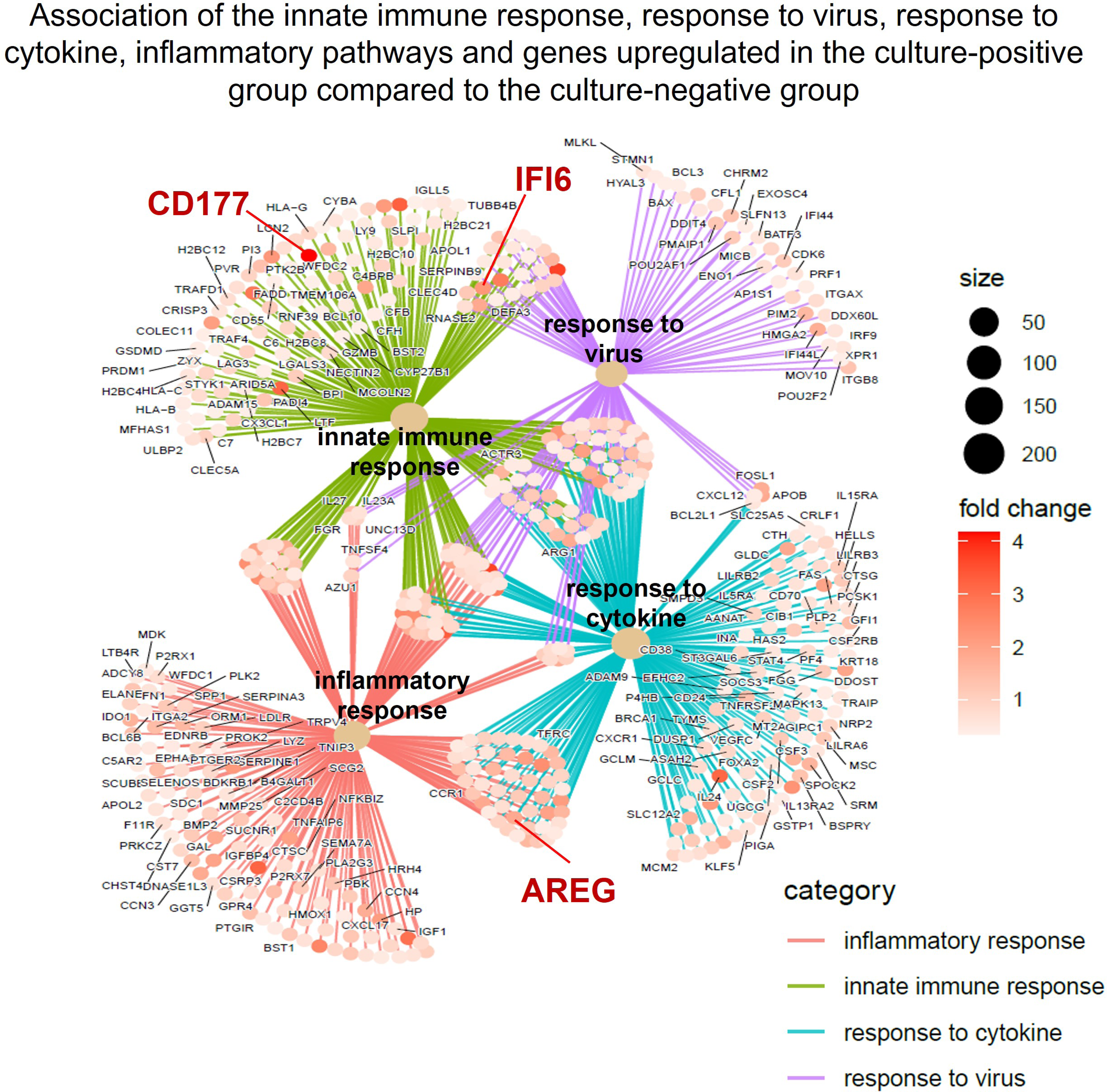
Transcriptomic analysis showing the association between the innate immune response, response to virus, response to cytokine, inflammatory pathways and genes upregulated in the culture-positive group versus the culture-negative group. The cnetplot illustrates the overlap of genes and their fold changes for selected activated pathways. Significant genes (p.adjust <0.05) that are annotated to the pathways are highlighted in red.

The differential expression (DE) results also revealed that a number of SARS-CoV-2 genes were significantly upregulated (FDR<0.01) in the culture-positive versus the culture-negative group including *nucleocapsid phosphoprotein* (log2 FC=8.4) and *ORF3a* (log2 FC=5.5) while the *surface/spike glycoprotein* encoding gene had a log2 FC of 5.3 and an FDR of 0.067 (Table S6). A visual inspection of the mapped SARS-CoV-2 reads revealed that those that mapped to the 5’ end of the genes were spliced with a portion mapping to the 5’ leader sequence of the genome. This suggests the reads originated from sub-genomic mRNA (sgRNA) rather than genomic RNA which is consistent with the active viral replication observed in the culture-positive group.

Finally, we evaluated whether any of the DE genes could act as biomarkers discriminating between lung culture-positive and negative-individuals. Logistic regression predictive modelling revealed that *GREM1* and *FGFBP1* were associated with a sensitivity and specificity above 90% (Figure S6). Future studies are warranted to determine if these lung-based biomarkers can predict patient culture status in blood samples.

## Discussion

The widely accepted view in severe acute COVID-19 is that resolution of the initial viral replication phase in the first week after symptom onset is followed by an effector or hyperinflammatory phase in the second and third week of illness, which is characterised by diffuse alveolar damage, thrombo-inflammation, and endotheliopathy (36). Indeed, the Infectious Disease Society of America (IDSA) recommends the use of remdesivir for only 5 days in patients with severe illness and not at all in mechanically ventilated patients (37). However, our results, based on post-mortem lung biopsies obtained using minimally invasive tissue sampling methods (MITS) shortly after death indicated that, in contradistinction to the URT where replication often ceases within ∼8 days from symptom onset, in the human lung virus is culturable in ∼40% of mechanically ventilated patients until death (median of 15 days and up to 4 weeks after symptom onset; see Figure 1 for the study overview). To ensure reproducibility of the lung biopsy procedure, histological analysis was performed to confirm that the tissue was derived from the lung only (to ensure that there was no contamination from other tissue or muscle, which would have been detected on histopathological analysis at the least to some extent). The upregulation of muscle-associated gene pathways may have been related to virus-associated myositis or ICU-associated myopathy. The culture-positive group in the Delta cohort had accelerated death and a higher proportion of secondary bacterial infection in the lung compared to the culture-negative group. This may be explained by the Delta variant being more transmissible (38), associated with enhanced replication, higher viral load (39) and greater immune escape (40) than the Beta variant.

Nasopharyngeal SARS-CoV-2 viral load (based on Ct value) neither at admission nor at death, was predictive of lung culture-positivity. SARS-CoV-2 culture-positivity in the lung of decedents was associated with attenuated pulmonary T-cell immunity and an exaggerated pro-inflammatory phenotype. Importantly, this was concurrent with, rather than sequential to the viral replication or viral culture-positive phase.

These findings challenge the traditional paradigm of an initial viral replicative phase in the first week of severe illness sequentially followed by an effector or inflammatory phase (36). Our data suggest that in ∼40% of ventilated patients, viral replication persisted until death (i.e. 3^rd^ and 4^th^ week of illness and a median of 15 days after symptom onset) compared to ∼2 to ∼8 days in the URT as outlined in several studies including a live virus human challenge study (11, 14–24). One outlier study reported culturing virus from the URT for up to 3 weeks after symptom onset (41). However, a large proportion of patients were immunocompromised, samples at diagnosis and follow-up were combined (skewing the results), a high proportion of participants were healthcare workers (re-infection may have been a confounder), and as the authors suggested a limitation was that the Vero cell line used was overtly permissive to infection compared to the human lung carcinoma cell line, H1299 ACE2, which is a biologically representative cell line (and one that we used). Another recent study showed that infectious virus production peaked in the human lung within 2 days, but this model used *ex vivo* agarose infused devascularised and explanted human lung slices, which are not representative of what is occurring in freshly harvested human lung (42). The culture-based findings in the afore-mentioned studies must be explicitly distinguished from studies that detected residual free viral genomic RNA (but not replicating virus) embedded in the respiratory tract tissue of patients that had severe disease for an extended period of time (13, 43, 44). Indeed, SARS-CoV-2 RNAs have been detected in patient tissue many months after recovery from acute infection (45–47). It was initially suggested that sgRNA (sub-genomic RNA; small strands of reversely transcribed RNA) could be used as a proxy to infer viral replication. However, several recent studies have indicated that it has poor predictive value as a proxy for viral replication (48, 49). Indeed, Stein et al (13) detected sgRNA in multiple post-mortem organ biopsies, including the brain, several months after symptom onset. Thus, the data presented in this manuscript is the first to do so conclusively and comprehensively using viral culture from lung tissue (the gold standard to detect viral replication) beyond two weeks after symptom onset.

We demonstrated active viral replication in the lungs of acutely ill ventilated patients for up to ∼4 weeks after symptom onset. This challenges the current practice of using antivirals like remdesivir for only 5 days and suggests that a longer duration of treatment may be required in critically ill patients. Furthermore, antivirals like remdesivir are not recommended for use by IDSA in mechanically ventilated patients (conditional recommendation) as they felt that such patients (often in the third week of their illness) are no longer in the viral replicative phase, and published controlled trial data showed no mortality benefit of remdesivir in such patients (37, 50). However, these studies demonstrated a group effect, and the analyses did not adjust for disease severity or the time from symptom onset to death in mechanically ventilated patients (51). Our data suggest that a significant number of patients may likely benefit from antivirals during mechanical ventilation. Indeed, several observational studies have found a survival benefit using remdesivir in mechanically ventilated patients, but this requires further clarification in appropriate trials (51–53). It is also possible that the very advanced immunopathology in some patients may render antivirals redundant. In a multivariate analysis we found no association between steroid usage and lung viral culture positivity - in fact, steroid usage was lower in the viral culture-positive group (and there was a trend to an inverse relationship in the multivariable analysis, and often culture positivity persisted beyond the 10 days of steroid usage.

The transcriptomic data suggested that, in a significant number of patients, the hyperinflammatory and viral replication phase occur concurrently in the 3^rd^ and 4^th^ week of illness, in contradistinction to the widely held view that these are sequential phases. Antiviral and selective proinflammatory responses were over-represented in the SARS-CoV-2 culture-positive compared with the culture-negative decedents, and we did not detect attenuated type 1 interferon responses at the site of disease compared with other reports (54–58). Three prior studies (one that enrolled 5 COVID-19 patients) evaluated transcriptomic lung responses in patients with severe COVID-19 versus healthy controls (54, 55, 59). These first level studies logically attempted to address the significance of transcriptomic changes specific to COVID-19 by using healthy controls or non-diseased parts of the lung from lung cancer patients. However, we specifically sought to compare culture-positive versus culture-negative groups (hitherto not undertaken) to dissect out pathways that facilitate permissiveness to ongoing viral replication.

We identified two lung-based biomarkers (*GREM1* and *FGFBP1*) that could predict culture-positivity. Although these are lung-specific biomarkers, this preliminary analysis in a limited number of samples suggests that in the future, RT-PCR of tracheal aspirates or blood (if they are concordant with lung findings), could potentially serve as biomarkers to identify and direct appropriate treatment protocols to culture-positive persons but further investigation is needed.

There are several limitations to our findings. Firstly, our findings are relevant to acute severe COVID-19 ARDS/pneumonia requiring mechanical ventilation and may not be applicable to milder forms of disease seen in hospitalised patients or chronic infection seen in immunocompromised patients. Second, we only studied patients with the Beta and Delta variants as these were the predominant variant at the time of the study. However, Omicron has also been associated with severe disease in several settings including the surge of severe COVID-19 unfolding in China. Third, we did not study a control group comprising severe ARDS due to other causes because our express aim was to investigate the presence and duration of viral replication in the LRT in severe COVID-19 disease. Fourth, the sample size limited our ability to make conclusions about several aspects. However, the highly resource intensive and demanding nature of the study limited our ability to recruit higher numbers of participants. Fifth, it could be suggested that there may have been sampling error and variability of the viral culture assay. However, the reproducibility of the viral culture technique using 6 samples across 2 separate runs had a low standard error, which was indicative of high reproducibility. Sixth, we did not compare the culture status of the lower respiratory tract in the ambulatory controls versus the decedents. This was due to ethical reasons and the potential risks of viral transmission to the medical and research staff during bronchoscopic procedures. Finally, the transcriptional signature and flow cytometric findings may have been affected by post-death sampling, but several detailed studies have shown (60) that most protein and RNA species are preserved and stable for several hours after death. Given that biopsies for the transcriptional studies were taken ∼2 hours after death, we feel they are broadly representative of the picture at the time of death. In summary, our data suggests that in COVID-19 disease there is considerable heterogeneity in the frequency and duration of viral replication in the upper versus the lower respiratory tract (i.e. lungs) beyond the 2^nd^ week of illness, and that in a significant proportion of seriously ill patients, persisting viral replication occurs concurrently and may drive an exaggerated proinflammatory response (higher than in culture-negative persons), rather than sequentially as it is widely believed. These findings have potential implications for the use of antiviral therapy in seriously ill patients with COVID-19 and suggest that better biomarkers are needed to identify patient phenotypes and subsets that might benefit from concurrent anti-inflammatory and antiviral therapy.

## Contributions

K.D, MT, S.J, A.P, A.E, M.D, M.N and S.M conceived and designed experiments. K.D, A.E, S.O, L.K, T.P, A.S, I.J, J.P, D.T, D.F, M.M, M.N and S.M arranged medical ethical approval, recruitment of study participants and collection of study material. MT, S.J, L.W, A.K, S.M, A.P, K.P, R.R, R.K, R.M, C.M, J.B, R.L, M.D, C vdM, A.R, M.H, V.B, S. M, C.W, and R.J performed the experiments. M.T, S.J, A.P and M.D set up experimental assays. A.S provided the cell line. K.D, M.T, S.J, L.W, A.K, L.S, S.M, A.P, K.P, R.R, R.L, M.H, V.B, S.M, R.J, and N.M analysed and interpreted data. K.D, M.T, L.S, and S.M wrote the manuscript with input from all listed authors.

## Declaration of interests

The authors have no competing interests.

## Data Availability

Individual participant data will be made available to researchers who provide a protocol that is approved by their respective human research ethics committee. All protocols will be reviewed and approved by the MITS consortium trial steering committee up to five years following publication. A data sharing agreement (DTA) will need to be concluded between the representatives of the requesting institution and the University of Cape Town Lung Institute. Data sharing requests should be directed to

## Acknowledgments

The authors would like to thank Arnold-Day C, Crowther M, Fernandes N, and Mitchell L from the Division of Critical Care, Department of Anaesthesia and Perioperative Medicine, University of Cape Town, South Africa for identifying and referring potential patients into the study.

The study was funded by the Bill & Melinda Gates Foundation (grant number INV-017282) and the South African Medical Research Council (grant number SHIP NCD 96756) with partial support from the Department of Science and Technology and National Research Foundation: South African Research Chair Initiative in Vaccine Preventable Diseases. The KD lab acknowledges funding from the SA MRC (RFA-EMU-02-2017), EDCTP (TMA-2015SF-1043, TMA-1051-TESAIII, TMA-CDF2015), UK Medical Research Council (MR/S03563X/1), NIH (CRDF-OISE-16-62105) and the Wellcome Trust (MR/S027777/1). This work was co-funded by The Wellcome Centre for Infectious Diseases Research in Africa is supported by core funding from the Wellcome Trust (230135/Z/16/Z) and the European Union’s Horizon Europe Research and Innovation Actions (101046041) for genomic surveillance.

The authors would like to thank the families of the deceased who gave us permission to conduct the study, which may advance our understanding of SARS-CoV-2 pathogenesis to inform future clinical management of respiratory pathogen pandemics.

## ONLINE DATA SUPPLEMENT

### Methods

#### Viral culture

The cell line was maintained in Roswell Parks Memorial medium (RPMI) containing 10% foetal bovine serum, 100 IU penicillin/streptomycin, 2 mM L-glutamine, 25 mM HEPES, 1× non-essential amino acids and 0.1 mg/mL sodium pyruvate (ThermoFisher, South Africa; Figure S1). The nasopharyngeal swabs in universal transport medium (UTM) were initially filtered through a 0.22µm filter prior to inoculation. The lung biopsy samples were placed in the well containing the cellular monolayer. The inoculated cultures were grown in a humidified 37°C incubator with 5% CO_2_ and cytopathic effect (CPE) and viral replication were monitored on days 1, 3, 6 and 9 by PCR. Viral culture positivity was defined as at least a 3-fold increase in viral load over time. Viral culture reproducibility was performed by a single observer with a total of 6 different viral culture experiments. Each viral culture was performed over a 6-day period with 3 sampling time points (days 1, 3 and 6) and the experiments were all plotted over the assay timepoints to enable line fitment between the data points. A R^2^ value of 0.94 (p=0.017) was obtained (1 being a perfect value), which indicated that the assay was highly reproducible.

#### Immunohistochemistry

Sections between 3-4µm thick were placed on adhesive slides and fixed at 37^°^C overnight. Heat induced epitope retrieval (HIER) time was set to 60 minutes to prevent tissue wash off and possible background staining. The antibodies (anti-CD3 [2GV6], anti-CD4 [SP35], anti-CD8 [SP57] and anti-CD68 [KP-1]; Roche USA) were incubated with the tissue sections for 30 minutes. After antibody and counter staining, slides were visualised using an Olympus BX41 microscope at 40x magnification.

#### SARS-CoV-2 whole genome sequencing

Total SARS-CoV-2 RNA was extracted from lung biopsy samples using the ChemagicTM 360 automated system (PerkinElmer, Inc, Waltham, MA) according to the chemagic Viral300 360 H96 drying prefilling VD200309.che protocol. Whole genome amplification and library preparation were performed using the Illumina COVIDSeq Test kit and protocol 1000000128490 v02 (Illumina, Inc., San Diego, CA), and executed on the Hamilton Next Generation StarLet (Hamilton Company). Whole genome amplification was achieved via multiplex polymerase chain reaction performed with the ARTIC V4.1 primers designed to generate 400-bp amplicons with an overlap of 70 bp that spans the 30 kb genome of SARS-CoV-2. Indexed paired-end libraries were normalized to 4 nM concentration, pooled, and denatured with 0.2 N sodium acetate. A 4pM pooled library was spiked with 1% PhiX Control v.3 adaptor-ligated library (Illumina, Inc., San Diego, CA) and sequenced using the MiSeq® Reagent Kit v2 (500 cycle) and sequenced on the MiSeq instrument (Illumina, Inc., San Diego, CA). The quality of sequencing reads was assessed using different tools including FastQC, Fastp, Fastv, Fastq_screen, and Fastx_toolkit. The resulting reads were analysed on Exatype (https://exatype.com/) for referenced-based genome assembly to identify minor and major variants. The assembled consensus sequences were analyzed using Nextclade Web (https://clades.nextstrain.org) for further quality control and clade assignment.

#### RNAseq

RNAseq was performed on lung post-mortem biopsy samples from 24 individuals which included 8 that were COVID culture-positive and 16 that were culture-negative.

Total RNA was extracted from lung biopsy samples using the RNeasy mini plus kit (Qiagen). Ribosomal depletion was performed, and libraries were prepared using the MGIEasy RNA Library Prep Set (Cat. No.: 1000006383, 1000006384, MGI, Shenzhen, China) as per manufacturer’s instructions. Sequencing was performed at the South African Medical Research Council Genomics Centre using DNA nanoball-based technology on the DNBSEQ-G400 (BGI, Shenzhen China) instrument generating 100 bp unstranded paired-end reads. The FastQC program [version 0.11.9; (1)], was used to assess read quality. The Spliced Transcripts Alignment to a Reference (STAR) software [version STAR_2.7.7a; (2)] was used to map reads consecutively to the Ensembl (3) human genome primary assembly (version GRCh38.109) and the SARS CoV-2 reference (ASM985889v3) with the quantMode and GeneCounts option selected to generate raw genewise read counts for each sample. A number of samples failed to pass QC due to a low number of mapped reads (< 2 million). A total of six culture-positive and five culture-negative samples were used in subsequent analysis.

The differential expression (DE) analysis was performed with the edgeR [version 3.38.4; (4)] Bioconductor (5) package. Briefly, raw counts were filtered to remove genes with low expression, normalized, and negative binomial generalized linear models were fitted. The likelihood ratio test was used to identify DE genes when comparing culture-positive to culture-negative samples.

A gene set enrichment analysis (GSEA) for Gene Ontology (Biological Process) was performed on the DE results ranked by fold change using the gseGO function, from the R clusterProfiler (ver: .4.4.4, PMID: 34557778) package.

## Supplementary results

**Figure S1.**
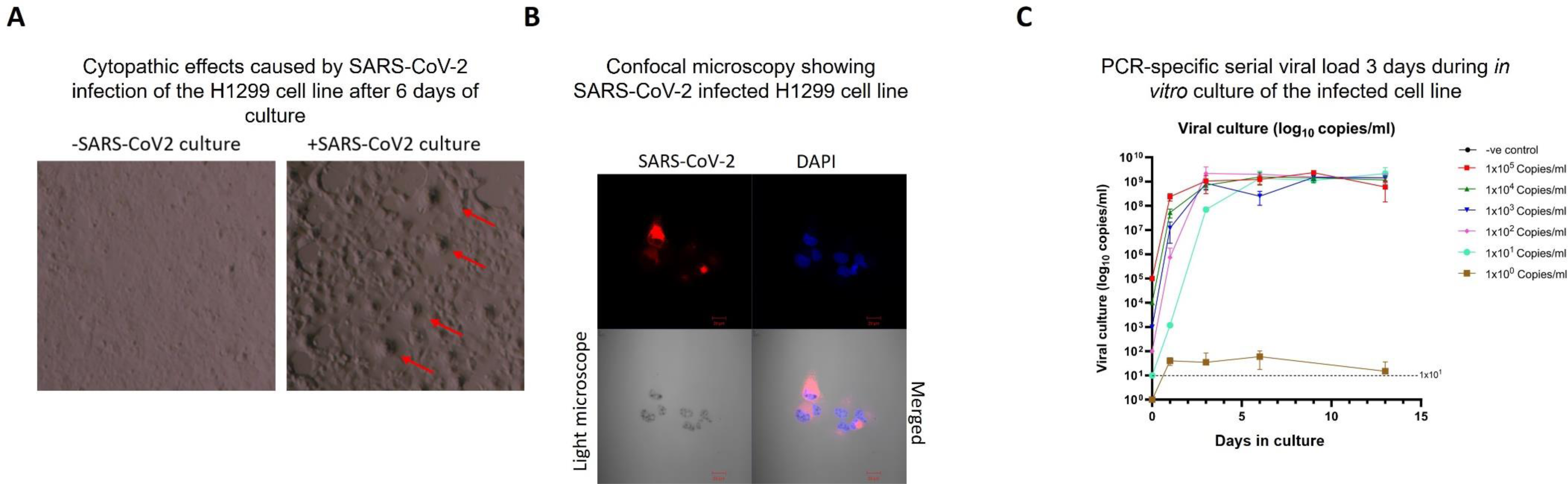
In vitro culture of SARS-CoV-2. (**A**) Light microscope images showing SARS-CoV-2 viral-induced cytopathic effects (red arrow). (**B**) Confocal microscopy showing SARS-CoV-2 (red) infecting the cell line. DAPI (blue) was used as the nuclear stain. (**C**) The limit of detection (LOD) for the PCR assay to detect replicating competent SARS-CoV-2. SARS-CoV-2 viral stock was diluted in 10-fold dilutions from 1×10^5^ to 1 copy/ml and co-cultured with confluent H1299 ACE2 cells in a 24-well plate for 9 days. Aliquots were analysed by PCR for viral load on days 1, 3, 6 and 9. The relative viral load (copies/ml) are shown. The dotted line represents the LOD for viral load (1×10^1^ copies/ml).

**Figure S2.**
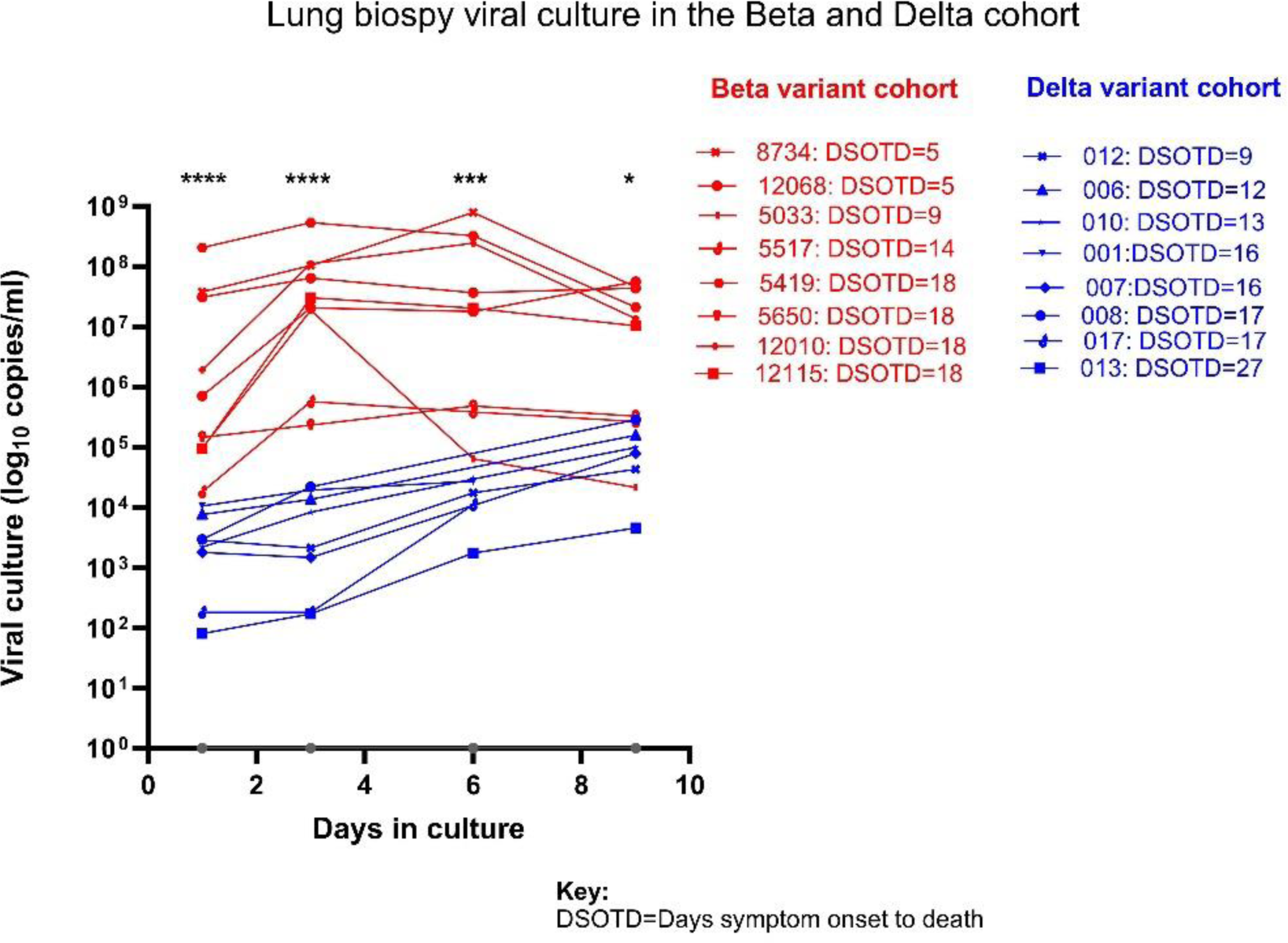
In vitro viral culture of lung biopsies from MV decedents. Description of a new SARS-CoV-2 human biophenotype that has ongoing viral replication in lung for up to 27 days post symptom onset. The lung cancer cell line, H1299 ACE2, was used to culture SARS-CoV-2. The lung biopsy samples were removed and placed in the well containing the cellular monolayer. The inoculated cultures were grown in a humidified 37°C incubator and viral replication were monitored on days 1, 3, 6 and 9 by PCR. DSOTD=days symptom onset to death.

**Figure S3.**
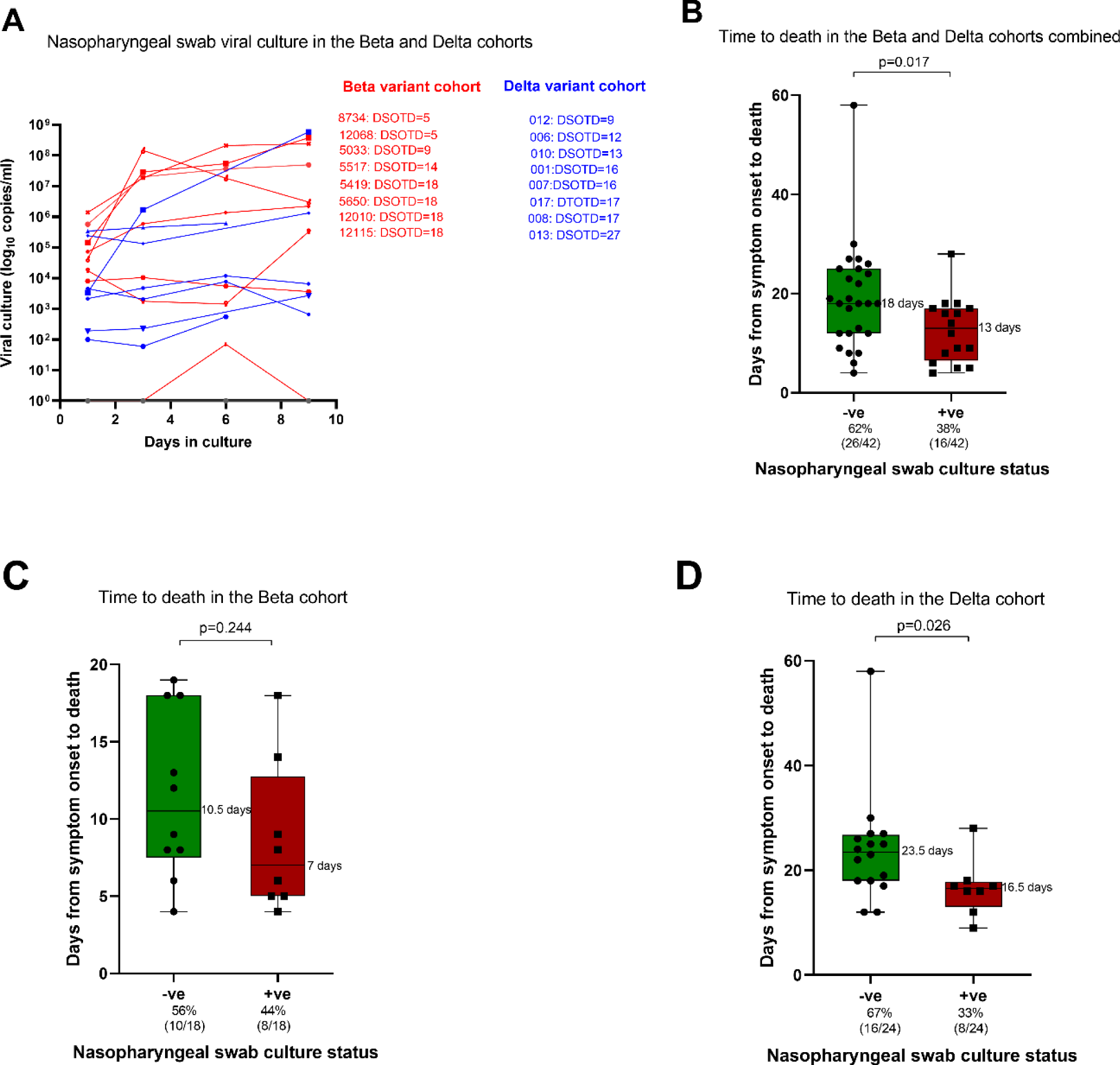
Previously ventilated decedents had active replicating virus in the URT for up to 27 days post symptom onset to death. (**A**) The lung cancer cell line, H1299 ACE2, was used to culture SARS-CoV-2. The nasopharyngeal swabs in universal transport medium were initially filtered through a 0.22µm filter prior to inoculation. The inoculated cultures were grown in a humidified 37°C incubator with 5% CO_2_ and cytopathic effect (CPE) and viral replication were monitored on days 1, 3, 6 and 9 by PCR. The days from symptom onset to death for the culture-negative (-ve; green) and culture-positive (+ve; red) groups are shown. The dotted lines represent the median days from symptom onset to death for the lung culture-positive (13 days) and lung culture-negative (18 days) decedents. The days from symptom onset to death for the culture-negative (-ve; green) and culture-positive (+ve; red) groups are shown for the Beta (**B**) and Delta (**C**) cohorts. The median days from symptom onset to death for the culture-positive (7 days for the Beta cohort and 16.5 days for the Delta cohort) and culture-negative (10.5 days for the Beta cohort and 23.5 days for the Delta cohort) participants are shown.

**Figure S4.**
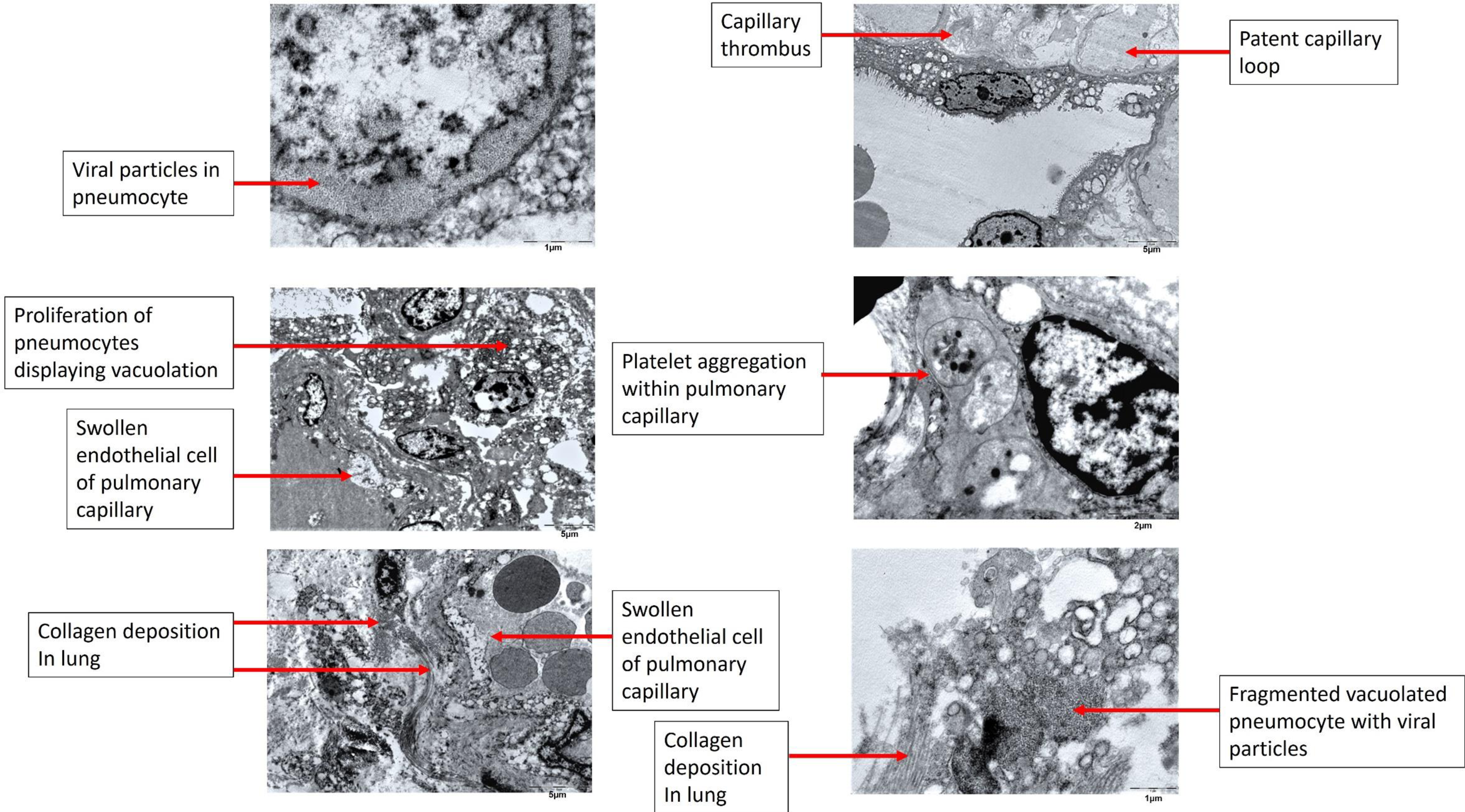
**Electron micrographs of key clinical features of lung abnormalities associated with acute COVID-19 disease.** The size of the representative scale bars are shown.

**Figure S5.**
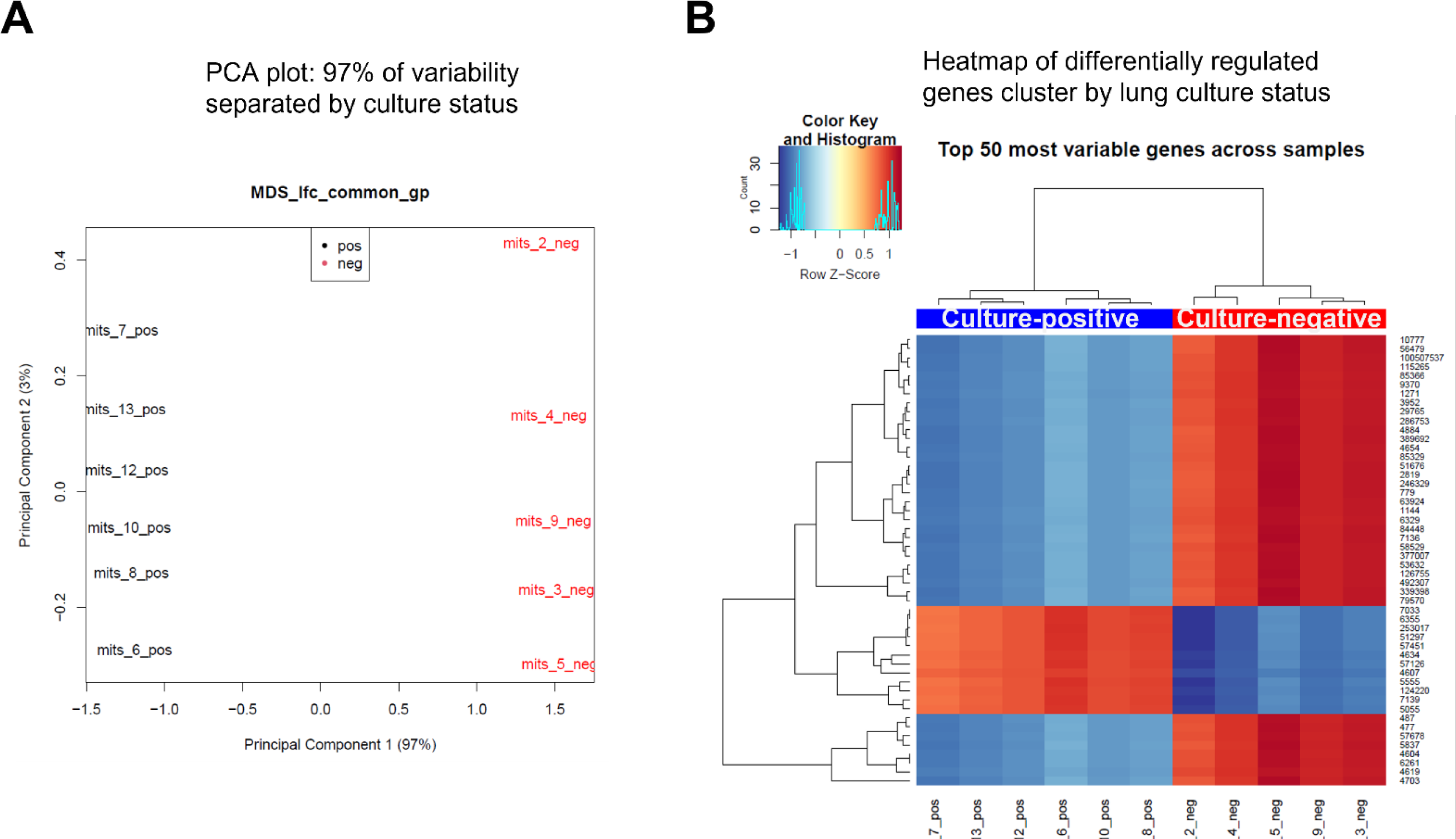
Gene expression profile in the culture-negative and culture-positive groups cluster into distinct groups. PCA plot **(A)** and heatmap **(B)** showing that distinct genes are differentially regulated in the culture-positive and culture-negative groups.

**Figure S6.**
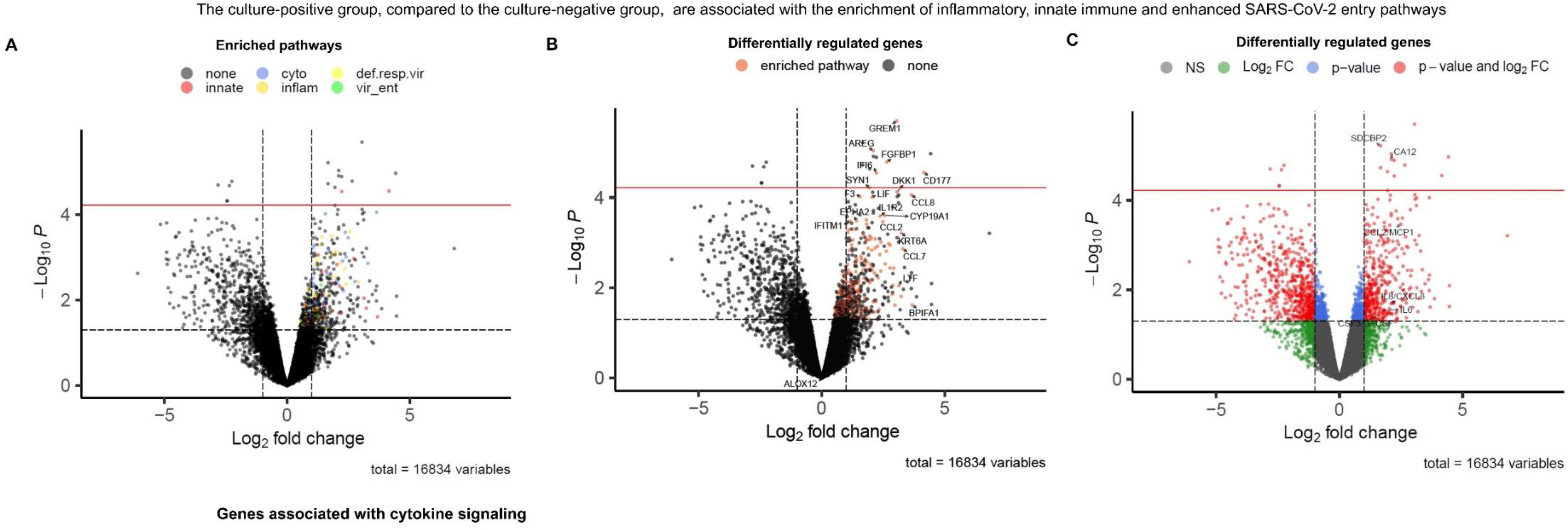
The culture-positive group expressed higher levels of genes associated with inflammatory, innate immunity and enhanced SARS-CoV-2 cellular entry pathways compared to the culture-negative cohort. Volcano plots showing the pathways (**A**) and individual genes (**B and C**) upregulated in the culture-positive versus the culture-negative cohort. Cyto = cytokine signalling, def. resp.virus = defence of respiratory virus, innate = innate immunology, inflame = inflammatory response, vir_ent=viral entry. The red line represents FDR<0.05 and the black dotted represents p<0.05.

**Table S1.**
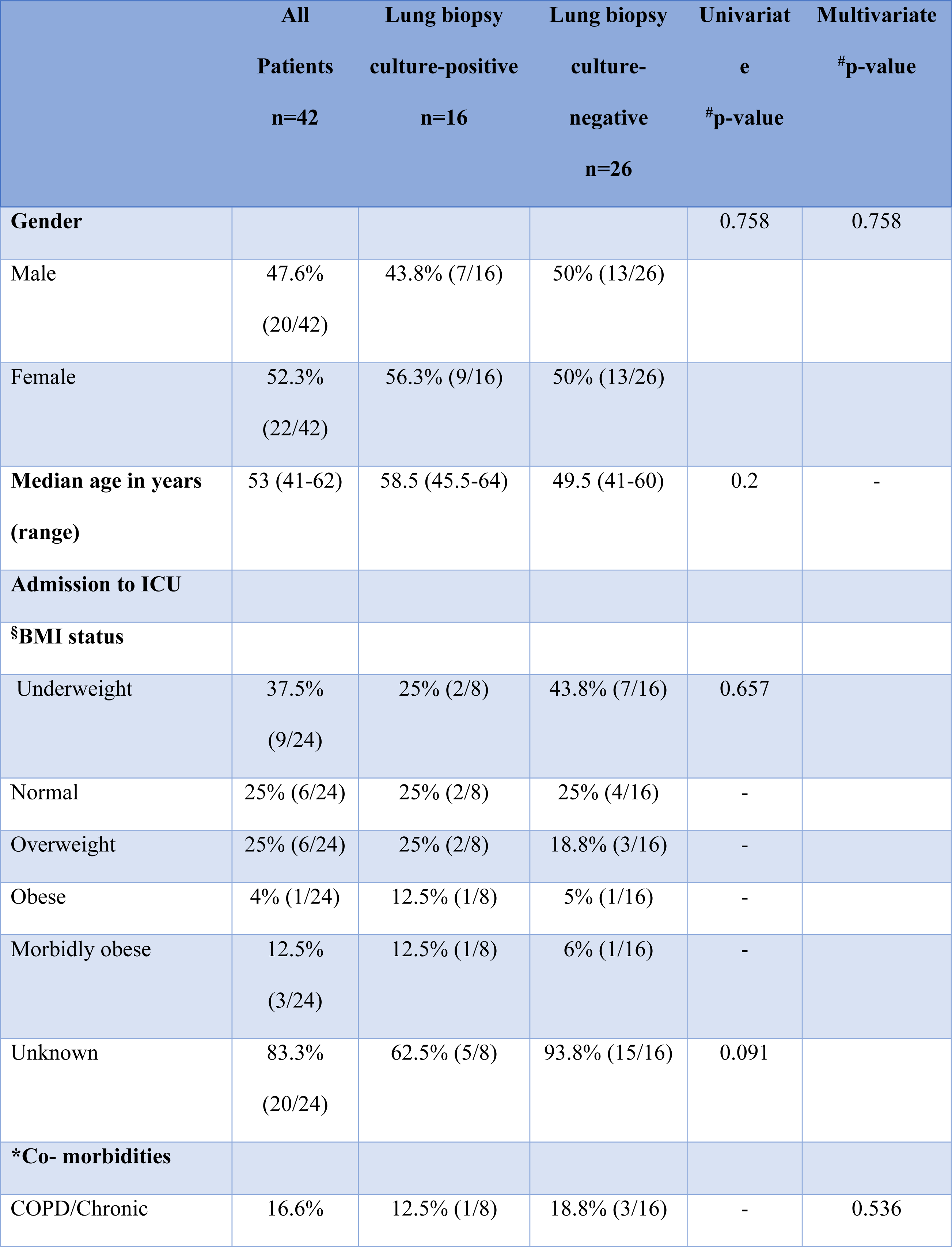
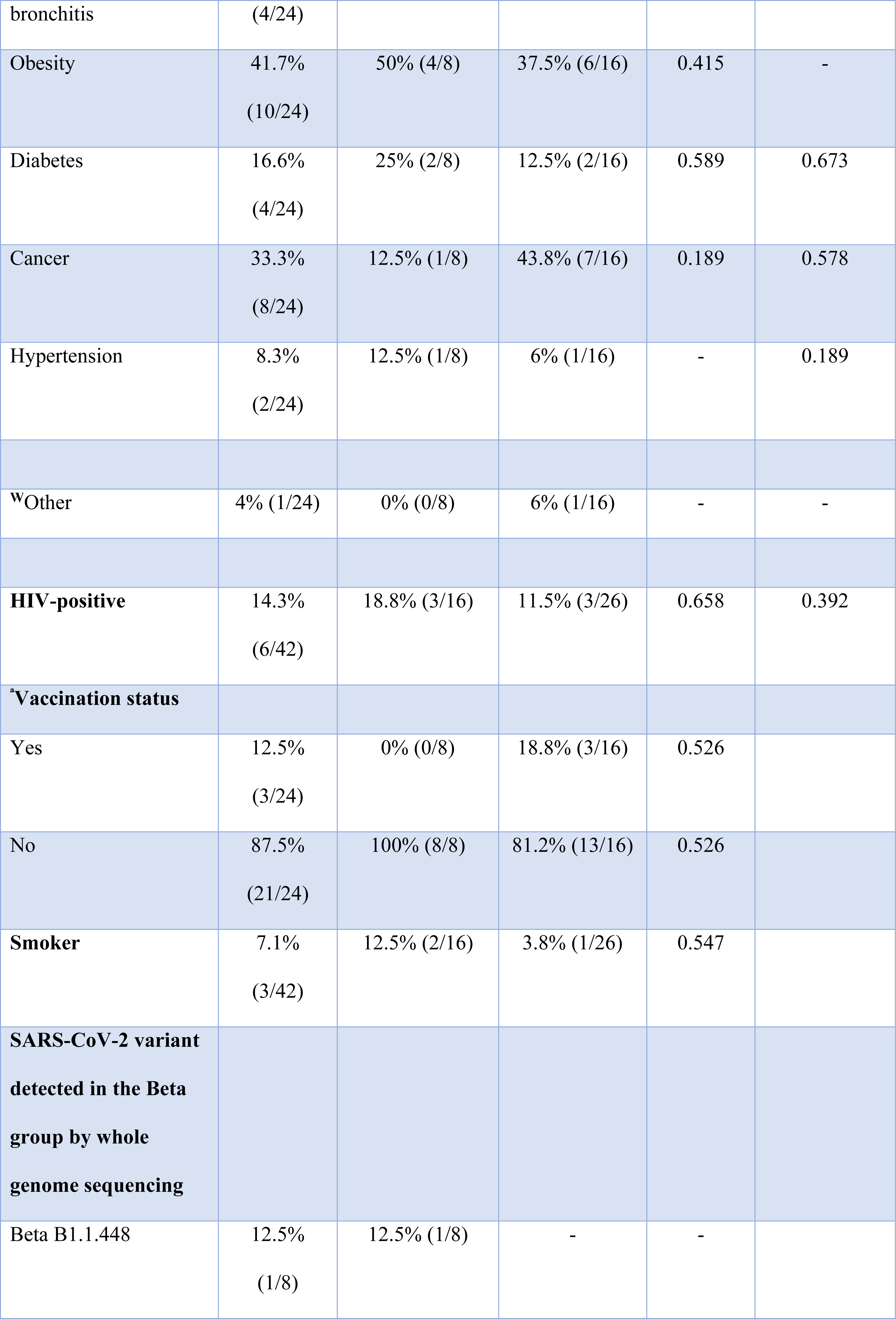
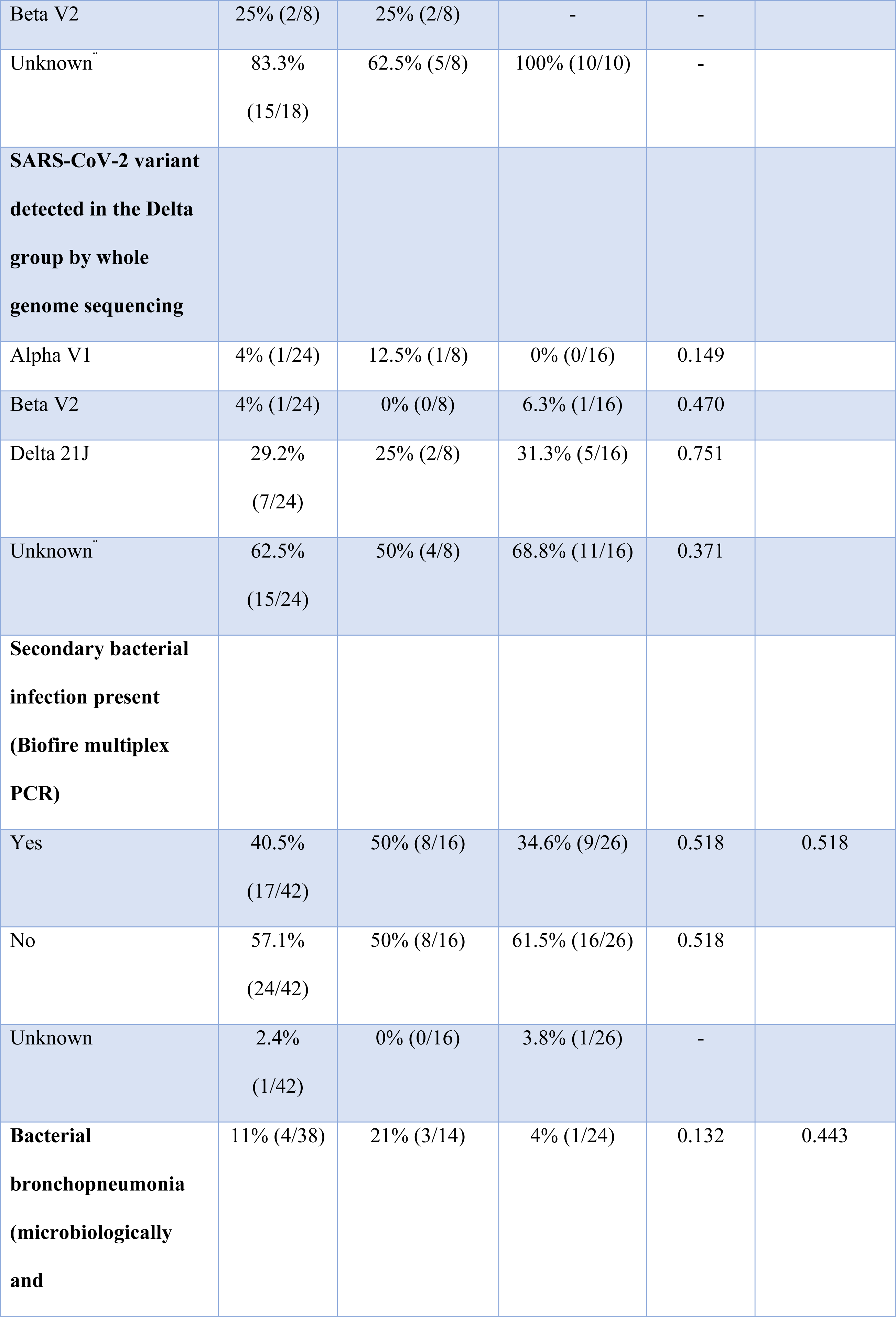
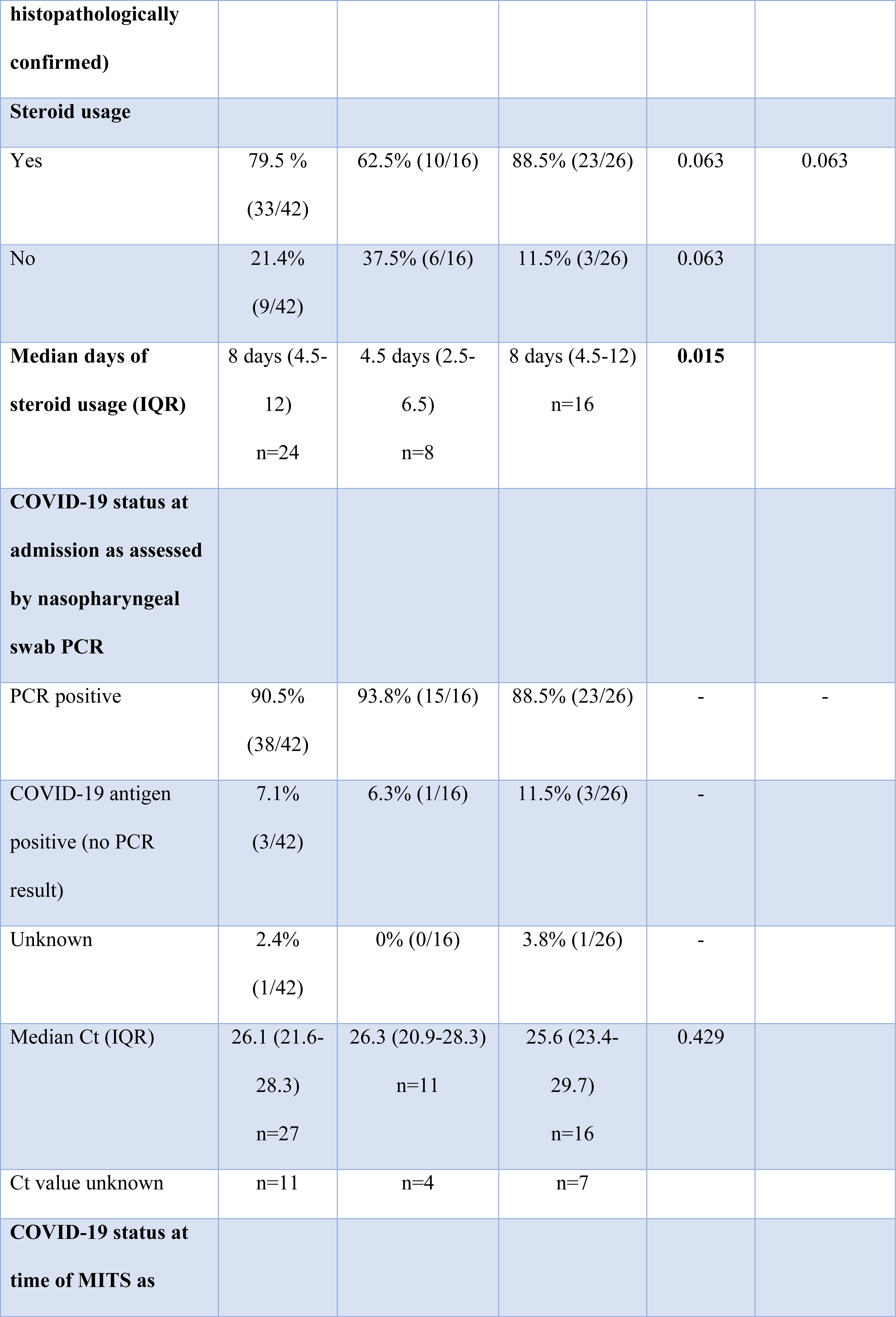
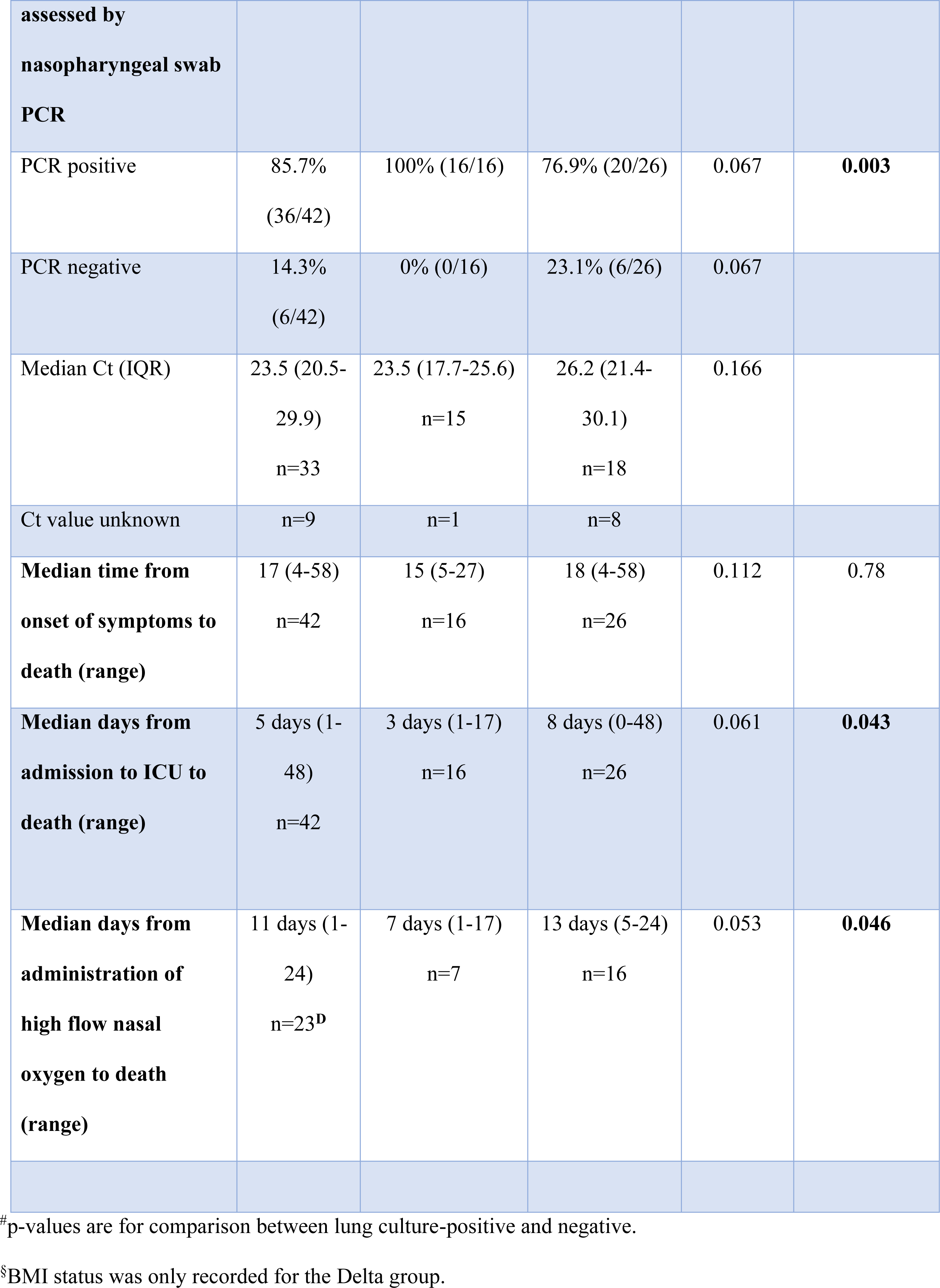
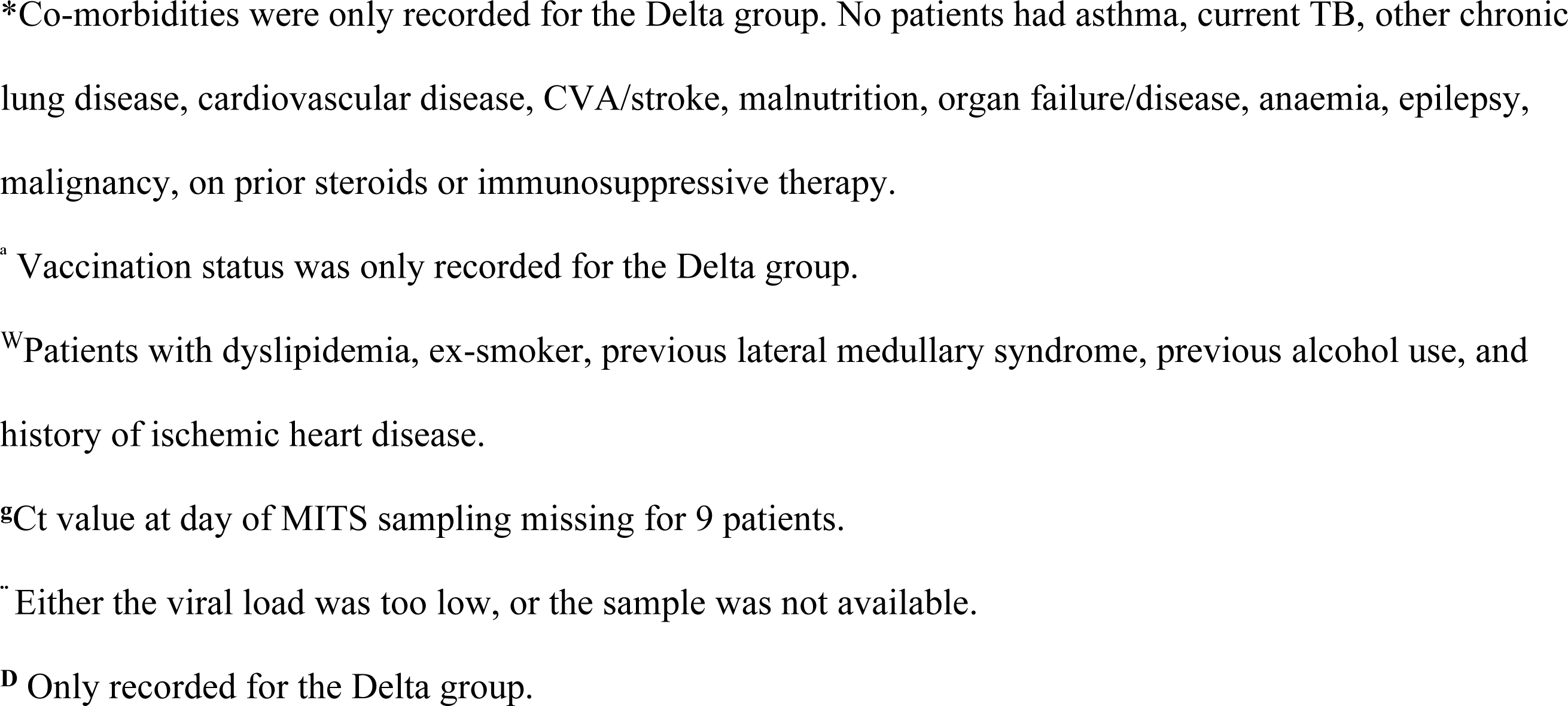
Demographic and clinical characteristics of the decedents.

**Table S2.** Histopathological data of the combined study cohort.

| **Histology Tissue** | **All patients**<br>**n=33\*** | **culture-**<br>**positive**<br>**patients**<br>**n=13** | **culture-**<br>**negative**<br>**patients**<br>**n=20** | **p-value** |
| --- | --- | --- | --- | --- |
| **LUNG** |  |  |  |  |
| Alveolar hyaline membranes | 24/33<br>(73%) | 8/13<br>(61.5%) | 16/20<br>(80%) | 0.425 |
| Interstitial oedema | 7/19 (37%) | 3/7<br>(43%) | 4/12 (33%) | - |
| Collapsed alveoli | 9/33 (27%) | 3/13<br>(23%) | 6/20 (30%) | - |
| Pneumocyte denudation/necrosis | 24/33<br>(73%) | 9/13<br>(69.2%) | 15/20<br>(75%) | - |
| Endothelial necrosis | 5/19 (26%) | 3/7<br>(43%) | 2/12 (17%) | 0.305 |
| Vascular neutrophil aggregate | 6/19 (32%) | 3/7<br>(43%) | 3/12 (25%) | 0.617 |
| Micro-thromboembolic | 11/33<br>(33%) | 5/13<br>(38.5%) | 6/20 (30%) | 0.714 |
| Pulmonary haemorrhage | 14/33<br>(42%) | 3/13<br>(23%) | 11/20<br>(55%) | 0.087 |
| Pulmonary endothelialitis | 5/19 (26%) | 2/7<br>(29%) | 3/12 (25%) | - |
| Organising hyaline membranes | 15/19<br>(79%) | 6/7<br>(86%) | 9/12 (75%) | - |
| Fibroblasts/myofibroblasts proliferation | 16/19<br>(84%) | 6/7<br>(86%) | 10/12<br>(83%) | - |
| Lymphocytic infiltration | 26/33<br>(79%) | 9/13<br>(69.2%) | 17/20<br>(85%) | 0.393 |
| Plasma cell infiltration | 4/19 (21%) | 1/7<br>(14%) | 3/12 (25%) | - |
| Type II pneumocytes proliferation | 30/33<br>(91%) | 10/13<br>(77%) | 20/20<br>(100%) | 0.052 |
| Atypical pneumocytes | 18/19<br>(95%) | 7/7<br>(100%) | 11/12<br>(92%) | - |
| Atypical pneumocyte cytomegaly | 25/33<br>(76%) | 9/13<br>(69%) | 16/20<br>(80%) | 0.681 |
| Atypical pneumocyte nucleomegaly | 21/33<br>(64%) | 7/13<br>(54%) | 14/20<br>(70%) | 0.465 |
| Atypical pneumocyte multinucleation | 19/33<br>(58%) | 6/13<br>(46%) | 13/20<br>(65%) | 0.472 |
| Atypical pneumocyte syncytia | 5/33 (15%) | 3/13<br>(23%) | 2/20 (10%) | 0.360 |
| Foamy pneumocytes | 19/19<br>(100%) | 7/7<br>(100%) | 12/12<br>(100%) | - |
| Arterial/arteriole thrombosis | 0/19 (0%) | 0/7 (0%) | 0/12 (0%) | - |
| Diffuse collagenous fibrosis | 0/19 (0%) | 0/7 (0%) | 0/12 (0%) | - |
| Subpleural/interstitial fibrosis | 7/19 (37%) | 2/7<br>(29%) | 5/12 (42%) | 0.657 |
| Honeycomb | 5/33 (15%) | 2/13<br>(15.4%) | 4/20 (20%) | - |
| Traction bronchiectasis | 0/19 (0%) | 0/7 (0%) | 0/12 (0%) | - |
| Squamous met | 1/19 (5%) | 1/7<br>(14%) | 0/12 (0%) | 0.368 |
| Intimal fibrosis | 0/19 (0%) | 0/7 (0%) | 0/12 (0%) | - |
| Medial hypertrophy | 0/19 (0%) | 0/7 (0%) | 0/12 (0%) | - |
| Organising pneumonia | 10/33<br>(30%) | 4/13<br>(31%) | 6/20 (30%) | - |
| Bronchopneumonia (neutrophilic infiltrate into the alveoli) | 10/33<br>(30%) | 5/13<br>(38.5%) | 5/20 (25%) | 0.461 |
| Intranuclear inclusions | 0/33 (0%) | 0/13<br>(0%) | 0/20 (0%) | - |
| Intracytoplasmic inclusions | 0/33 (0%) | 0/13<br>(0%) | 0/20 (0%) | - |
| Megakaryocytes | 22/33<br>(68%) | 8/13<br>(71%) | 14/20<br>(70%) | 0.714 |
| Lung haemophagocytosis (increased alveolar macrophages) | 17/33<br>(52%) | 3/13<br>(23%) | 14/20<br>(70%) | **0.013** |
| Lung siderophages | 6/33 (18%) | 2/13<br>(15.4%) | 4/20 (20%) | - |
| Necrotising granulomas | 1/33 (3%) | 1/13<br>(0%) | 0/20 (7.7%) | 0.625 |
| Non-necrotising granulomas | 0/33 (0%) | 0/13(0%) | 0/20 (0%) | - |
| **HEART** |  |  |  |  |
| Isolated myocyte necrosis | 6/33 (18%) | 1/13<br>(14%) | 5/20 (25%) | 0.364 |
| Cardiac ischaemia | 8/17 (47%) | 3/7<br>(43%) | 5/10 (50%) | - |
| Cardiac thrombi | 1/33 (3%) | 0/13<br>(0%) | 1/20 (5%) | - |
| Card capillary neutrophil<br>margination | 0/17 (0%) | 0/7 (0%) | 0/10 (0%) | - |
| RBC fragment | 0/17 (0%) | 0/7 (0%) | 0/10 (0%) | - |
| Lipofuscin | 17/17<br>(100%) | 7/7<br>(100%) | 10/10<br>(100%) | - |
| Enlarged nuclei | 23/33<br>(70%) | 10/13<br>(77%) | 13/20<br>(65%) | 0.467 |
| Cardiac interstitial fibrosis | 20/33<br>(61%) | 4/13<br>(31%) | 16/20<br>(80%) | **0.001** |
| Heart endothelialitis | 1/33 (3%) | 0/13<br>(0%) | 1/20 (10%) | - |
| Cardiac neutrophils | 0/33 (0%) | 0/13<br>(0%) | 0/20 (0%) | - |
| Cardiac lymphocytes | 5/33 (15%) | 1/13<br>(7.7%) | 4/20 (20%) | 0.625 |
| Cardiac histiocytes | 2/33 (6%) | 1/13<br>(7.7%) | 1/20 (5%) | - |
| Cardiac eosinophils | 1/33 (3%) | 0/13<br>(0%) | 1/20 (5%) | - |
| Cardiac interstitial oedema | 4/33 (12%) | 0/13<br>(0%) | 4/20 (20%) | 0.136 |
| **LIVER** |  |  |  |  |
| Increase Kupffer | 16/17<br>(94%) | 7/7<br>(100%) | 9/10 (90%) | - |
| Foamy Kupffer | 4/17 (24%) | 1/7<br>(14%) | 3/10 (30%) | 0.603 |
| Liver haemophagocytosis | 9/33 (27%) | 4/13<br>(31%) | 5/20 (25%) | - |
| Liver siderophages | 1/33 (3%) | 0/13<br>(0%) | 1/20 (5%) | - |
| Liver necrosis | 7/17 (41%) | 3/7<br>(43%) | 4/10 (40%) | - |
| Spotty necrosis zone 1 | 0/17 (0%) | 0/7 (0%) | 0/10 (0%) | - |
| Spotty necrosis zone 2 | 2/16 (13%) | 1/6<br>(17%) | 1/10 (10%) | - |
| Spotty necrosis zone 3 | 2/17 (12%) | 1/7<br>(14%) | 1/10 (10%) | - |
| Confluent necrosis zone 1 | 0/17 (0%) | 0/7 (0%) | 0/10 (0%) | - |
| Confluent necrosis zone 2 | 1/17 (6%) | 0/7 (0%) | 1/10 (10%) | - |
| Confluent necrosis zone 3 | 4/17 (24%) | 1/7<br>(14%) | 3/10 (30%) | 0.603 |
| Micro steatosis | 14/17<br>(82%) | 6/7<br>(86%) | 8/10 (80%) | - |
| Macro steatosis | 13/17<br>(76%) | 6/7<br>(86%) | 7/10 (70%) | - |
| Cholestasis | 8/33 (24%) | 3/13<br>(23%) | 5/20 (25%) | - |
| Liver inflammation | 9/17 (53%) | 3/7<br>(43%) | 6/10 (60%) | 0.637 |
| Liver regeneration | 16/17<br>(94%) | 7/7<br>(100%) | 9/10 (90%) | - |
| Liver congestion | 9/33 (%) | 8/13<br>(61.5%) | 11/20<br>(55%) | - |
| Liver viral inclusions | 0/17 (0%) | 0/7 (0%) | 0/10 (0%) | - |
| Liver fibrosis F1 | 2/17 (12%) | 1/7<br>(14%) | 1/10 (10%) | - |
| Liver fibrosis F2 | 0/17 (0%) | 0/7 (0%) | 0/10 (0%) | - |
| Liver fibrosis F3 | 1/17 (6%) | 0/7 (0%) | 1/10 (10%) | - |
| Liver fibrosis F4 | 1/17 (6%) | 0/7 (0%) | 1/10 (10%) | - |
| Primary sclerosing cholangitis | 0/17 (0%) | 0/7 (0%) | 0/10 (0%) | - |
| Primary biliary cholangitis | 0/17 (0%) | 0/7 (0%) | 0/10 (0%) | - |
| Extramedullary haematopoiesis<br>(EMH) | 0/17 (0%) | 0/7 (0%) | 0/10 (0%) | - |
| **KIDNEY** |  |  |  |  |
| Glom capillary dilatation | 10/11<br>(91%) | 5/5<br>(100%) | 5/6 (83%) | - |
| Glom thrombus | 4/11 (36%) | 0/5 (0%) | 4/6 (67%) | 0.061 |
| Glom sclerosis | 3/11 (27%) | 0/5 (0%) | 3/6 (50%) | 0.182 |
| Increase mesangium | 7/11 (64%) | 3/5<br>(60%) | 4/6 (67%) | - |
| Membrane thickening | 0/11 (0%) | 0/5 (0%) | 0/6 (0%) | - |
| Acute kidney injury | 4/11 (36%) | 0/5 (0%) | 4/6 (67%) | 0.061 |
| Tubular casts | 6/11 (55%) | 2/5<br>(40%) | 4/6 (67%) | 0.242 |
| Isometric vacuole | 0/11 (0%) | 0/5 (0%) | 0/6 (0%) | - |
| Kidney interstitial lymphocytes | 1/11 (9%) | 1/5<br>(20%) | 0/6 (0%) | - |
| Kidney interstitial plasma | 1/11 (9%) | 1/5<br>(20%) | 0/6 (0%) | - |
| Kidney interstitial fibrosis | 2/11 (18%) | 0/5 (0%) | 2/6 (33%) | 0.455 |
| Kidney interstitial haem | 0/11 (0%) | 0/5 (0%) | 0/6 (0%) | - |
| Kidney arteriosclerosis | 4/11 (36%) | 1/5<br>(20%) | 3/6 (50%) | 0.546 |
| **ADIPOSE** |  |  |  |  |
| Fat necrosis | 10/21<br>(48%) | 6/8<br>(75%) | 4/13 (31%) | 0.082 |
| Fat fibrin | 0/21 (0%) | 0/8 (0%) | 0/13 (0%) | - |
| Fat unremarked | 11/21 (52) | 2/8<br>(25%) | 9/13 (69%) | 0.082 |

\*Histopathology was not performed on all the biopsy samples.

**Table S3.** Electron microscopy data of the Delta cohort.

| **Histology Tissue** | **All patients**<br>**n=17\*** | **culture-**<br>**positive**<br>**patients**<br>**n=7** | **culture-**<br>**negative**<br>**patients**<br>**n=10** | **p-value** |
| --- | --- | --- | --- | --- |
| **LUNG** |  |  |  |  |
| Pneumocyte vacuolation | 13/17<br>(76%) | 6/7<br>(86%) | 7/10 (70%) | 0.603 |
| Endothelial vacuolation | 3/17 (18%) | 2/7<br>(29%) | 1/10 (10%) | 0.537 |
| Endothelial swelling | 13/17<br>(76%) | 6/7<br>(86%) | 7/10 (70%) | 0.603 |
| Activated capillary monocytes | 1/17 (6%) | 0/7 (0%) | 1/10 (10%) | - |
| Pneumocyte detachment | 15/17<br>(88%) | 7/7<br>(100%) | 8/10 (80%) | 0.485 |
| Collagen deposits | 11/16<br>(69%) | 5/6<br>(83%) | 6/10 (60%) | 0.588 |
| **HEART** |  |  |  |  |
| Mitochondrial hypoxic changes | 12/12<br>(100%) | 6/6<br>(100%) | 6/6 (100%) | - |
| Myocyte atrophy and wrinkling | 11/11<br>(100%) | 6/6<br>(100%) | 5/5 (100%) | - |
| Lipofuscin | 7/11 (64%) | 5/6<br>(83%) | 2/5 (40%) | 0.242 |
| Swollen endothelial | 10/11<br>(91%) | 6/6<br>(100%) | 4/5 (80%) | - |
| Fragmented red blood cells | 4/11 (36%) | 4/6<br>(57%) | 0/5 (0%) | 0.061 |
| Myofibrillar disruption | 5/11 (45%) | 4/6<br>(57%) | 1/5 (20%) | 0.242 |
| **LIVER** |  |  |  |  |
| Swollen/hypoxic mitochondria | 15/15<br>(100%) | 6/6<br>(100%) | 9/9 (100%) | - |
| Increased Kupffer | 2/15 (13%) | 1/6<br>(17%) | 1/9 (11%) | - |
| Steatosis | 7/15 (47%) | 2/6<br>(33%) | 5/9 (56%) | 0.398 |
| Haemophagocytosis | 10/15<br>(67%) | 4/6<br>(57%) | 6/9 (67%) | - |
| Detached of endothelial cells | 7/15 (47%) | 3/6<br>(50%) | 4/9 (44%) | - |
| Lipidized stellate cells | 7/15 (47%) | 3/6<br>(50%) | 4/9 (44%) | - |
| EM liver cholestasis | 2/14 (14%) | 1/6<br>(17%) | 1/8 (13%) | - |
| **ADIPOSE** |  |  |  |  |
| Normal mitochondria | 3/14 (21%) | 0/6 (0%) | 3/8 (38%) | 0.209 |
| Slightly enlarged mitochondria | 5/14 (36%) | 4/6<br>(57%) | 1/8 (13%) | 0.091 |
| Enlarged mitochondria | 6/14 (43%) | 2/6<br>(33%) | 4/8 (50%) | 0.627 |
| Swollen endothelial cells present | 9/11 (82%) | 4/4<br>(100%) | 5/7 (71%) | 0.491 |

\*EM was only performed on biopsy samples in the Delta cohort.

**Table S4.**
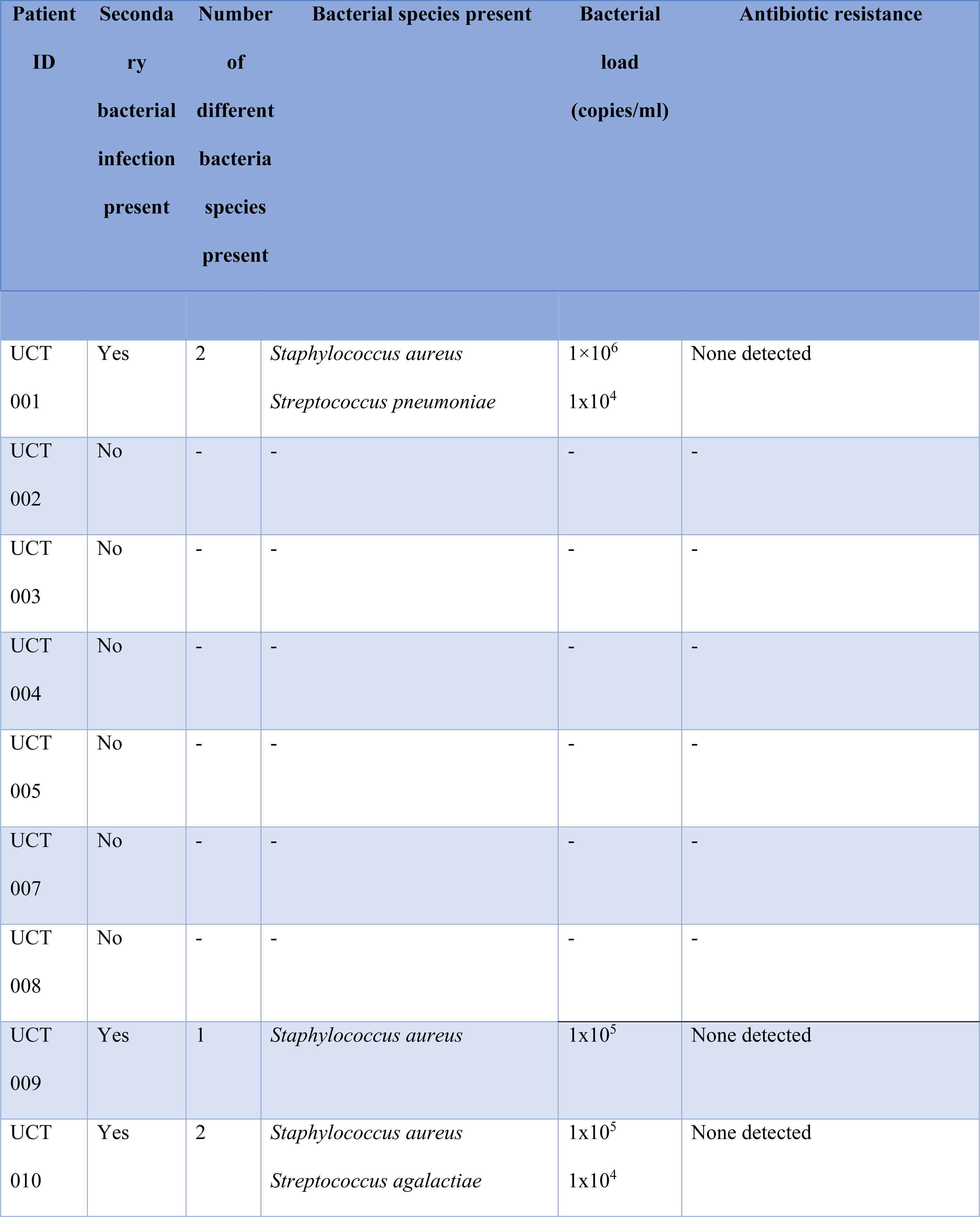
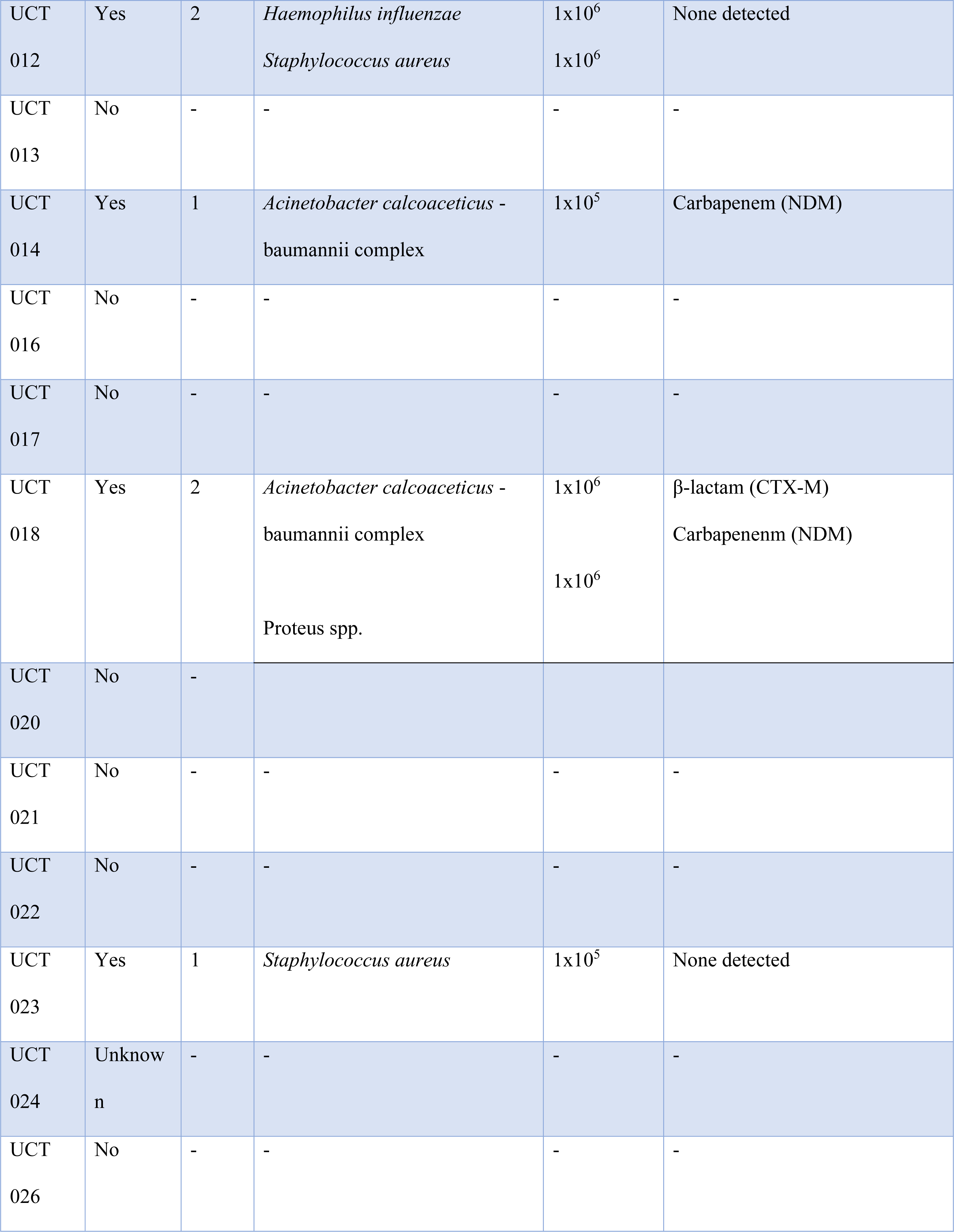
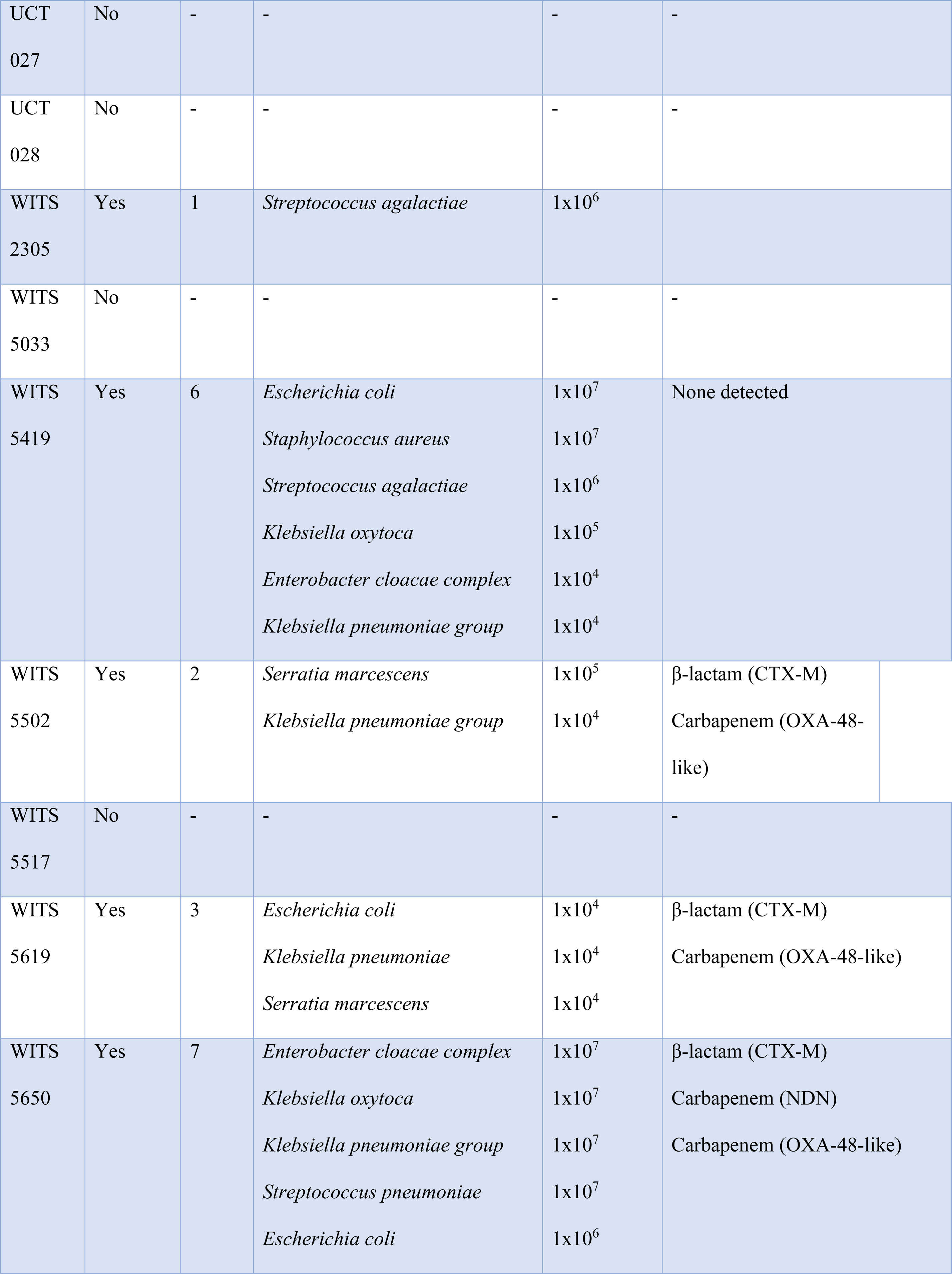
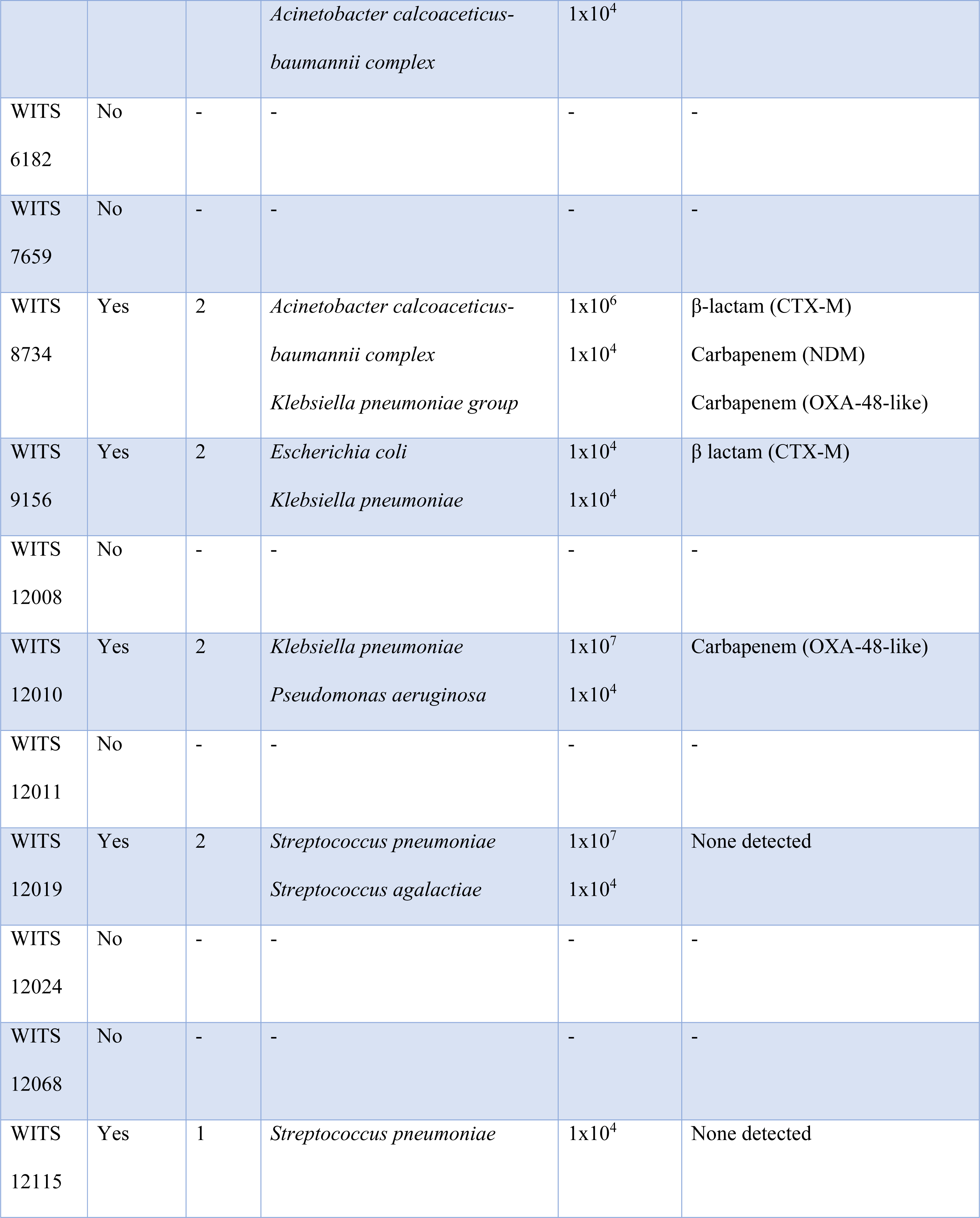
Bacteria and antibiotic resistance profile detected from the lung biopsies of the decedents using multiplex PCR (Biofire).

| Patient ID | Secondary bacterial infection present | Number of different bacteria species present | Bacterial species present | Bacterial load (copies/ml) | Antibiotic resistance |
| --- | --- | --- | --- | --- | --- |
| UCT 001 | Yes | 2 | *Staphylococcus aureus*<br>*Streptococcus pneumoniae* | $1 \times 10^6$<br>$1 \times 10^4$ | None detected |
| UCT 002 | No | - | - | - | - |
| UCT 003 | No | - | - | - | - |
| UCT 004 | No | - | - | - | - |
| UCT 005 | No | - | - | - | - |
| UCT 007 | No | - | - | - | - |
| UCT 008 | No | - | - | - | - |
| UCT 009 | Yes | 1 | *Staphylococcus aureus* | $1 \times 10^5$ | None detected |
| UCT 010 | Yes | 2 | *Staphylococcus aureus*<br>*Streptococcus agalactiae* | $1 \times 10^5$<br>$1 \times 10^4$ | None detected |
| UCT<br>012 | Yes | 2 | *Haemophilus influenzae*<br>*Staphylococcus aureus* | 1x10<sup>6</sup><br>1x10<sup>6</sup> | None detected |
| UCT<br>013 | No | - | - | - | - |
| UCT<br>014 | Yes | 1 | *Acinetobacter calcoaceticus* -<br>baumannii complex | 1x10<sup>5</sup> | Carbapenem (NDM) |
| UCT<br>016 | No | - | - | - | - |
| UCT<br>017 | No | - | - | - | - |
| UCT<br>018 | Yes | 2 | *Acinetobacter calcoaceticus* -<br>baumannii complex<br>Proteus spp. | 1x10<sup>6</sup><br>1x10<sup>6</sup> | β-lactam (CTX-M)<br>Carbapenem (NDM) |
| UCT<br>020 | No | - |  |  |  |
| UCT<br>021 | No | - | - | - | - |
| UCT<br>022 | No | - | - | - | - |
| UCT<br>023 | Yes | 1 | *Staphylococcus aureus* | 1x10<sup>5</sup> | None detected |
| UCT<br>024 | Unknown | - | - | - | - |
| UCT<br>026 | No | - | - | - | - |
| UCT<br>027 | No | - | - | - | - |
| UCT<br>028 | No | - | - | - | - |
| WITS<br>2305 | Yes | 1 | *Streptococcus agalactiae* | 1x10<sup>6</sup> |  |
| WITS<br>5033 | No | - | - | - | - |
| WITS<br>5419 | Yes | 6 | *Escherichia coli*<br>*Staphylococcus aureus*<br>*Streptococcus agalactiae*<br>*Klebsiella oxytoca*<br>*Enterobacter cloacae complex*<br>*Klebsiella pneumoniae group* | 1x10<sup>7</sup><br>1x10<sup>7</sup><br>1x10<sup>6</sup><br>1x10<sup>5</sup><br>1x10<sup>4</sup><br>1x10<sup>4</sup> | None detected |
| WITS<br>5502 | Yes | 2 | *Serratia marcescens*<br>*Klebsiella pneumoniae group* | 1x10<sup>5</sup><br>1x10<sup>4</sup> | β-lactam (CTX-M)<br>Carbapenem (OXA-48-like) |
| WITS<br>5517 | No | - | - | - | - |
| WITS<br>5619 | Yes | 3 | *Escherichia coli*<br>*Klebsiella pneumoniae*<br>*Serratia marcescens* | 1x10<sup>4</sup><br>1x10<sup>4</sup><br>1x10<sup>4</sup> | β-lactam (CTX-M)<br>Carbapenem (OXA-48-like) |
| WITS<br>5650 | Yes | 7 | *Enterobacter cloacae complex*<br>*Klebsiella oxytoca*<br>*Klebsiella pneumoniae group*<br>*Streptococcus pneumoniae*<br>*Escherichia coli* | 1x10<sup>7</sup><br>1x10<sup>7</sup><br>1x10<sup>7</sup><br>1x10<sup>7</sup><br>1x10<sup>6</sup> | β-lactam (CTX-M)<br>Carbapenem (NDN)<br>Carbapenem (OXA-48-like) |
|  |  |  | *Acinetobacter calcoaceticus-baumannii complex* | 1x10<sup>4</sup> |  |
| WITS 6182 | No | - | - | - | - |
| WITS 7659 | No | - | - | - | - |
| WITS 8734 | Yes | 2 | *Acinetobacter calcoaceticus-baumannii complex*<br>*Klebsiella pneumoniae group* | 1x10<sup>6</sup><br>1x10<sup>4</sup> | β-lactam (CTX-M)<br>Carbapenem (NDM)<br>Carbapenem (OXA-48-like) |
| WITS 9156 | Yes | 2 | *Escherichia coli*<br>*Klebsiella pneumoniae* | 1x10<sup>4</sup><br>1x10<sup>4</sup> | β lactam (CTX-M) |
| WITS 12008 | No | - | - | - | - |
| WITS 12010 | Yes | 2 | *Klebsiella pneumoniae*<br>*Pseudomonas aeruginosa* | 1x10<sup>7</sup><br>1x10<sup>4</sup> | Carbapenem (OXA-48-like) |
| WITS 12011 | No | - | - | - | - |
| WITS 12019 | Yes | 2 | *Streptococcus pneumoniae*<br>*Streptococcus agalactiae* | 1x10<sup>7</sup><br>1x10<sup>4</sup> | None detected |
| WITS 12024 | No | - | - | - | - |
| WITS 12068 | No | - | - | - | - |
| WITS 12115 | Yes | 1 | *Streptococcus pneumoniae* | 1x10<sup>4</sup> | None detected |

**Table S5.** Whole genome sequencing results of the viral variants from the study cohort. Only 6 culture-negative and 6 culture-positive samples were available for sequencing because either the viral load was too low (Ct>30) or the sample was not available. It was therefore impossible to perform comparative analysis between the 2 groups because of the small sample number. Data was analysed and sequences aligned using the Stanford University Coronavirus Antiviral and Resistance Database (available at: https://covdb.stanford.edu/).

| Patient ID* | Source of sample | Variant | Lineage | Sub-lineage | Gene | Unique mutations (found in <0.01% of genomes) | Comments | Ref |
| --- | --- | --- | --- | --- | --- | --- | --- | --- |
| WITS 1210 | Lung culture supernatant | Beta | 20B | B.1.1 | - | None |  |  |
| WITS 12068 | Lung culture supernatant | Beta | 20H | B.1.3.5.1 | - | None |  |  |
| WITS 12115 | Lung culture supernatant | Beta | 20H | B.1.351 | - | None |  |  |
| UCT 001 | NPS | Alpha | 20I | B1.1.7 | - | None |  |  |
| UCT 004 | NPS | Beta | 20H | B1.351 |  |  |  |  |
| UCT 028 | NPS | Delta | 21J | AY6 | - | None |  |  |
| UCT 016 | NPS | Delta | 21J | AY91 |  |  |  |  |
| UCT 008 | NPS | Delta | 21J | AY32 | Papain-like protease (PLpro) | C55C*W<br>E113EAGV<br>E115ED<br>C118C*W |  |  |
|  |  |  |  |  | Helicase (nsp13) | P593DE<br>R594X |  |  |
|  |  |  |  |  | mRNA capping | K433KN<br>P443PAST |  |  |
|  |  |  |  |  | protein<br>(nsp 14) | K469KMRT<br>A471APST |  |  |
|  |  |  |  |  | Membrane (M) | S211R<br>S212*DEHQ<br>Y<br>S213X |  |  |
| **UCT 012** | NPS | Delta | 21J | AY32 | - | None |  |  |
| **UCT 018** | NPS | Delta | 21J | B1.617.2 | Main protease (PLpro) | A191T | A191T is a reported resistance mutation against 3C-like protease inhibitor, i.e. nirmatrelvir. | (6) |
|  |  |  |  |  | RdRP | H650N |  |  |
|  |  |  |  |  | Spike | V16VFIL<br>K77E |  |  |
|  |  |  |  |  | Nucleocapsid | S255A |  |  |
| **UCT 022** | NPS | Delta | 21J | AY32 | - | Sequence missing |  |  |
| **UCT 024** | NPS | Delta | 21J | AY6 | Main protease (PLpro) | N1454NIST |  |  |
|  |  |  |  |  | Helicase<br>(nsp13) | F587L<br>T588TIR |  |  |
|  |  |  |  |  | mRNA<br>capping<br>protein<br>(nsp 14) | M72X<br>R485DE<br>H486X<br>H487X |  |  |
|  |  |  |  |  | Accessor<br>y protein<br>(ORF3a) | K21R<br>D22D*EHQY<br>L94LV |  |  |

**Table S6.** Transcriptomic analysis showing the DE genes in the culture-positive cohort relative to the culture-negative cohort. Genes in black text are human specific. Genes in red text are SARS-CoV-2 specific.

| ENZ_ID | Abbreviation | Gene name | logFC | logCPM | p value | FDR |
| --- | --- | --- | --- | --- | --- | --- |
| 43740569 | ORF3a | ORF3a protein | 8.35 | -0.02 | 3,23E-07 | **0.005** |
| 43740575 | N | Nucleocapsid phosphoprotein | 5.5 | 2.14 | 1,09E-06 | **0.009** |
| 26585 | GREM1 | Gremlin 1, DAN family BMP antagonist | 3.1 | 5.43 | 2,85E-06 | **0.016** |
| 100271927 | RASA4B | RAS p21 protein activator 4B | -2.6 | 3.21 | 4,27E-06 | **0.018** |
| 27111 | SDCBP2 | Syndecan binding protein 2 | 1.67 | 3.38 | 8,53E-06 | **0.029** |
| 374 | AREG | Amphiregulin | 2.11 | 5 | 1,10E-05 | **0.031** |
| 771 | CA12 | Carbonic anhydrase 12 | 2.11 | 5.34 | 1,57E-05 | **0.034** |
| 5055 | SERPINB2 | Serpin family B member 2 | 4.43 | 2.5 | 1,81E-05 | **0.034** |
| 134285 | TMEM171 | Transmembrane protein 171 | 2.22 | -0.55 | 1,82E-05 | **0.034** |
| 338328 | GPIHBP1 | Glycosylphosphatidylinositol anchored high density lipoprotein binding protein 1 | -2.25 | 2.63 | 2,16E-05 | **0.035** |
| 9982 | FGFBP1 | Fibroblast growth factor binding protein 1 | 2.65 | 2.07 | 2,44E-05 | **0.035** |
| 80320 | SP6 | Sp6 transcription factor | 1.67 | 1.8 | 2,53E-05 | **0.035** |
| 1847 | DUSP5 | Dual specificity phosphatase 5 | 1.95 | 5.27 | 2,87E-05 | **0.035** |
| 2537 | IFI6 | Interferon alpha inducible protein 6 | 2.22 | 6.62 | 2,95E-05 | **0.035** |
| 57126 | CD177 | CD177 molecule | 4.15 | 5.53 | 4,26E-05 | **0.047** |
| 4915 | NTRK2 | Neurotrophic receptor tyrosine kinase 2 | -2.453 | 4.7 | 5,63E-05 | 0.059 |
| 43740568 | S spike | Surface glycoprotein/spike glycoprotein | 5.32 | 2.06 | 6,84E-05 | 0.067 |
| 23105 | FSTL4 | Follistatin like 4 | -1.25 | 0.8 | 7,35E-05 | 0.068 |
| 6853 | SYN1 | Synapsin I | 1.95 | 2.28 | 8,13E-05 | 0.071 |
| 2827 | GPR3 | G protein-coupled receptor 3 | 2.07 | 0.53 | 9,07E-05 | 0.076 |

**Table S7A.** Pathways upregulated in the culture-positive cohort relative to the culture-negative cohort.

| GO.ID | Pathway | Total number of genes associated with the pathway | Enrichment score | NES | p value | FDR |
| --- | --- | --- | --- | --- | --- | --- |
| GO:0045087 | Innate immune response | 644 | 0,462041652 | 2,23077102 | 3,19E-26 | 1,95E-22 |
| GO:0009617 | Response to bacterium | 502 | 0,436586937 | 2,063633461 | 1,46E-16 | 2,30E-13 |
| GO:0006954 | Inflammatory response | 666 | 0,397117204 | 1,910719799 | 1,84E-16 | 2,30E-13 |
| GO:0009615 | Response to virus | 337 | 0,482017466 | 2,192737214 | 4,76E-16 | 4,15E-13 |
| GO:0034097 | Response to cytokine | 784 | 0,378837973 | 1,853887449 | 4,57E-16 | 4,15E-13 |
| GO:0042742 | Defence response to bacterium | 187 | 0,57065003 | 2,468893487 | 2,87E-15 | 1,95E-12 |
| GO:0097529 | Myeloid leukocyte migration | 197 | 0,560938069 | 2,429088511 | 7,89E-15 | 4,82E-12 |
| GO:0071345 | Cellular response to cytokine stimulus | 700 | 0,379135852 | 1,832624826 | 1,29E-14 | 7,19E-12 |
| GO:0060326 | Cell chemotaxis | 260 | 0,505415546 | 2,244480472 | 3,87E-14 | 1,69E-11 |
| GO:0030595 | Leukocyte chemotaxis | 199 | 0,543874673 | 2,355877738 | 7,37E-14 | 2,81E-11 |
| GO:1903047 | Mitotic cell cycle process | 679 | 0,37365134 | 1,802308085 | 9,44E-14 | 3,39E-11 |
| GO:0042330 | Taxis | 521 | 0,406071568 | 1,918906559 | 1,71E-13 | 5,80E-11 |
| GO:0050900 | Leukocyte migration | 334 | 0,461740475 | 2,094951262 | 1,90E-13 | 6,01E-11 |
| GO:0006935 | Chemotaxis | 519 | 0,406436107 | 1,934745764 | 1,97E-13 | 6,01E-11 |
| GO:0000280 | Nuclear division | 380 | 0,433965649 | 1,999760733 | 8,85E-13 | 2,35E-10 |
| GO:0019221 | Cytokine-mediated signalling pathway | 401 | 0,424428923 | 1,972848247 | 2,15E-12 | 5,46E-10 |
| GO:0007059 | Chromosome segregation | 305 | 0,457222273 | 2,071558403 | 2,72E-12 | 6,65E-10 |
| GO:0051301 | Cell division | 573 | 0,383696506 | 1,825281074 | 3,41E-12 | 7,87E-10 |
| GO:2000147 | Positive regulation of cell motility | 507 | 0,386349508 | 1,825320536 | 7,25E-12 | 1,53E-09 |
| GO:0002237 | Response to molecule of bacterial origin | 294 | 0,453670477 | 2,036098196 | 1,01E-11 | 2,05E-09 |

**Table S7B.** Other pathways of interest upregulated in the culture-positive cohort relative to the culture-negative cohort.

| GO.ID | Pathway | Total number of genes associated with the pathway | Enrichment score | NES | FDR |
| --- | --- | --- | --- | --- | --- |
| GO:04657 | IL-17 signalling pathway | 81 | 0,635892 | 2.380892 | 1,53E-07 |
| GO:0002456 | T-cell-mediated immunity | 96 | 0,478752825 | 1,850434828 | 1,1E-03 |
| GO:0002825 | Regulation of T-helper 1 type immune response | 25 | 0,576145234 | 1,920646442 | 1,2E-3 |
| GO:0035710 | CD4-positive, alpha-beta T cell activation | 89 | 0,470392439 | 1,801972021 | 2,1E-03 |
| GO:0035743 | CD4-positive, alpha-beta T cell cytokine production | 15 | 0,774743052 | 2,055120611 | 3E-03 |
| GO:0002726 | Positive regulation of T cell cytokine production | 24 | 0,683190625 | 2,000311209 | 3,4E-03 |
| GO:0042110 | T cell activation | 448 | 0,294698908 | 1,379497641 | 3,5E-03 |
| GO:2000514 | Regulation of CD4-positive, alpha-beta T cell activation | 56 | 0,519437379 | 1,853870959 | 5,8-03 |
| GO:0002711 | Positive regulation of T cell mediated immunity | 49 | 0,543959461 | 1,869533223 | 6,7E-3 |
| GO:0042088 | Th1 immune response | 41 | 0,576145234 | 1,920646442 | 6,7E-03 |
| GO:0002709 | Regulation of T-cell mediated immunity | 76 | 0,686930834 | 2,050809398 | 7,3E-03 |
| GO:0002369 | T-cell cytokine production | 34 | 0,610833263 | 1,94794025 | 9,1E-3 |
| GO:0002724 | Regulation of T cell cytokine production | 34 | 0,610833263 | 1,94794025 | 9,1E-03 |
| GO:0050868 | Negative regulation of T cell activation | 109 | 0,413862853 | 1,63976229 | 1,1E-02 |

**Table S7C.** Genes associated with regulatory pathways of were not up- or downregulated in the culture-positive cohort relative to the culture-negative cohort.

| ENZ ID | Gene | LogFC | p-value | FDR |
| --- | --- | --- | --- | --- |
| 5133 | PD-1 | -0,580384672 | 0,352556526 | 0,875617669 |
| 64115 | VISTA | 0,195588418 | 0,601343382 | 0,941688131 |
| 84868 | TIM-3 | 0,243920799 | 0,568480759 | 0,936258857 |
| 1493 | CTLA-4 | 0,100226405 | 0,868488938 | 0,987403587 |
| 940 | CD28 | -0,515589978 | 0,192859859 | 0,773804865 |
| 945 | CD33 | -0,047967186 | 0,910411079 | 0,991670954 |
| 3559 | CD25 | -0,703108858 | 0,154569866 | 0,728675358 |
| 50943 | FOXP3 | -0,918764993 | 0,158003877 | 0,732211968 |

## Notes

### Competing Interest Statement

The authors have declared no competing interest.

### Funding Statement

The study was funded by the South African Medical Research Council.

### Author Declarations

Ethical approval was obtained from the Human Research Ethics Committee (HREC) of the University of Cape Town (HREC approval number 866/2020) and University of Witwatersrand (HREC approval number M200313). Biosafety approvals were obtained from the Faculty Biosafety Committee of the University of Cape Town (IBC008-2021).

### Summary of Updates

Good day. We have added how we ensured reproducibility during the lung biopsy procedure. Added discussion on the implications of steroid and antibiotic usage. The genes that were upregulated in the immune pathways have now been labelled in the transcripomic figure. Figure 5C now appears as its own figure for clarity. Magnification now appears on the EM and H&E images.

